# Mapping disordered eating symptoms in the ABCD Study through the Eating Disorder Symptom Index

**DOI:** 10.64898/2026.11.10.26367686

**Authors:** Carolina Makowski, Devansh Bansal, Brooke Moorman, Kimberly Garcia, Laura Ziemer, Janosch Linkersdörfer, Deanna Barch, Kelsey Hagan, Kyung E. Rhee, Shana Adise

**Affiliations:** Department of Psychiatry, University of California San Diego, La Jolla, California, USA; Department of Nutritional Sciences, University of Georgia, Athens, GA, USA; Department of Psychiatry, Virginia Commonwealth University, Richmond, Virginia, USA; Center for Human Development, University of California San Diego, La Jolla, California, USA; Center for Multimodal Imaging and Genetics, J. Craig Venter Institute, La Jolla, CA, USA; Department of Psychological and Brain Sciences, Washington University, St. Louis, MO, USA; Department of Pediatrics, University of California San Diego, La Jolla, California, USA; Department of Pediatrics, Division of Endocrinology, Diabetes and Metabolism, Children’s Hospital Los Angeles, Los Angeles, California, USA

## Abstract

Eating disorders (EDs) are serious psychiatric conditions that typically onset during adolescence and are often marked by a chronic and severe illness course. Capturing ED symptoms as they emerge is critical for charting risk markers and associated outcomes to inform early intervention and treatment efforts. The Adolescent Brain Cognitive Development^SM^ (ABCD^®^) Study is one such key study that could accelerate these efforts, but has been limited to date by a lack of clear, clinically informed definitions capturing youth at risk for ED-related concerns. The current manuscript aims to address this gap by defining ED symptom indices (ED_SI_) with a reproducible framework developed in collaboration with ED researchers and clinicians to identify youth with patterns of ED symptoms and behaviors that approximate subthreshold and diagnostic-like presentations. We describe item-level definitions and decision rules used to construct the ED_SI_, which include symptom indices of anorexia nervosa, bulimia nervosa, binge eating disorder, and other specified feeding or eating disorders. We then compare youth meeting ED_SI_ criteria with those identified by existing ED categorical definitions in ABCD Study data releases. By providing transparent definitions and analytical tools for identifying youth with potentially clinically meaningful ED behaviors, the ED_SI_ framework aims to facilitate more reproducible and interpretable research related to EDs using the ABCD Study dataset, while also providing a structure to examine the emergence, persistence, and change of ED-like symptoms longitudinally across adolescence and into young adulthood in future research.

## INTRODUCTION

Eating disorders (EDs) are severe psychiatric illnesses that typically emerge in adolescence (Galmiche et al., 2019; Uhlhaas et al., 2023). They are characterized by persistent disruptions in eating behaviors and related cognitive processes that impair food consumption. EDs often result in marked physical and/or psychosocial impairment (American Psychiatric Association, 2022), with extreme cases leading to mortality (Attia & Walsh, 2025; Hambleton et al., 2022; Krug et al., 2025). Globally, approximately 22% of children and adolescents report symptoms of disordered eating, with 1.2% of males and 5.7% of females aged 11-19 receiving a clinical diagnosis (López-Gil et al., 2023; Volpe et al., 2016). Mid-to-late adolescence (∼age 15.5) represents the peak period for ED symptom onset (Hudson et al., 2007; Solmi et al., 2022). This is a critical time for intervention, as untreated EDs are associated with increased risk of persistent illness and impairment. Consequently, it is imperative to catch emerging ED symptoms early to facilitate faster referral for treatment (Attia & Guarda, 2022; Kalindjian et al., 2022; Mills et al., 2024).

The gold standard for diagnosing EDs is through comprehensive, clinician-administered interviews. These interviews assess cognitive and behavioral symptom frequency, duration, weight history, compensatory behaviors, and psychosocial impairment secondary to the ED (American Psychiatric Association, 2022; Forbush et al., 2020). However, accessible and clinically-informed self-report questionnaires have also been developed for broader, non-specialist use when clinician-administered interviews are not feasible or practical (Kaufman et al., 1997; Luce & Crowther, 1999; Morgan et al., 1999), such as in large population-level and longitudinal cohort studies. One such questionnaire is the Kiddie Schedule for Affective Disorders and Schizophrenia - Computerized Version (KSADS-COMP). The KSADS-COMP is a guided, self-administered, computerized assessment that aims to mirror the *Diagnostic and Statistical Manual of Mental Disorders*, 5th Edition (*DSM*-5) (American Psychiatric Association, 2022). It has been widely used and validated for youth ages 6-18 and their caregivers (Kaufman et al., 1997; Townsend et al., 2020) and assesses weight distress, weight-control behaviors (e.g., caloric restriction, purging), binge eating, and related psychopathology.

It is important to acknowledge that current ED categories derived from the KSADS-COMP and similar assessments mirror *DSM-5-like* symptoms rather than clinical diagnoses. Additionally, branching logic and scoring algorithms are proprietary, posing challenges for rigor and reproducibility (Martin et al., 2026). For example, in the ABCD Study, the branching logic for the KSADS-COMP ED module is not openly available, making it difficult for users to determine the exact criteria used to include participants in specific categories. Further, the naming of categories generated by proprietary software are often misinterpreted as diagnostic labels in youth samples, as evidenced by papers using these categories to compare with epidemiological estimates of eating disorders (Murray et al., 2022; Rozzell et al., 2019). Despite these limitations, non-clinical assessment questionnaires remain highly valuable for identifying ED behaviors and symptoms in large, non-clinical samples (Hail et al., 2024; Lauth et al., 2010; Marques et al., 2022; Townsend et al., 2020).

Therefore, the current manuscript examines the extent to which ED classifications derived from questionnaire-based assessments such as the KSADS-COMP align with *DSM-5* diagnostic criteria. Using data from the Adolescent Brain Cognitive Development (ABCD) Study, a large, diverse longitudinal cohort study in which caregivers completed the ED module of the KSADS-COMP annually (and youth biannually), we developed symptom-level indicators that apply transparent, *DSM-5*-informed logic to the available KSADS-COMP items. The goal is not to establish clinical diagnoses, but to provide researchers with a reproducible framework for identifying youth with patterns of ED symptoms and behaviors that approximate *DSM-5* presentations, including potentially clinically meaningful subthreshold presentations. While these eating disorder symptom indices (ED_SI_) were developed in collaboration with clinicians who work with and/or treat youth with EDs and researchers with expertise in ED assessment, the numbers of youth meeting criteria in this manuscript are for descriptive purposes only. Together, these new ED symptom indices (ED_SI_) were developed with clear branching logic to promote reproducible and transparent research of ED-like behaviors in the ABCD Study.

We first describe the item-level definitions and decision rules used to construct the ED_SI_. We then compare the youth identified by the ED_SI_ with those identified using existing KSADS-COMP-derived classifications, examining the extent of agreement and discrepancy between the two approaches. These comparisons are intended to characterize general differences between classification methods, which may be due to proprietary methods. Finally, because the ABCD Study administered the KSADS-COMP repeatedly across development, the ED_SI_ provides a framework for examining the emergence, persistence, and change in ED-like symptoms longitudinally. By providing transparent definitions and reproducible analytic tools, this work aims to facilitate more rigorous and interpretable research on ED symptoms using questionnaires such as the KSADS-COMP. Collectively, this manuscript provides a roadmap for developing the ED_SI_.

## METHODS

### KSADS-COMP Assessment

The KSADS-COMP is a self-administered computerized questionnaire completed by caregivers and youth. Caregivers were given the option to complete this questionnaire in English or Spanish, whereas only the English version was available to youth; it was completed in person or via Zoom (Karcher & Barch, 2020). In the 2-and 4-year follow-up visits, the KSADS-COMP questions were read to youth by a research assistant. From the 6-year follow-up onward, youth completed questionnaires independently, but were monitored by a research assistant. While caregivers completed this questionnaire annually, youth reports were collected only biannually beginning at the 2-year follow-up assessment.

### Variable naming KSADS-COMP

Item-level response data were collected using KSADS-COMP’s web interface, scored using proprietary algorithms, and subsequently transferred to the ABCD Study as item-level, symptom-level, and “diagnosis” variables. Note that the KSADS-COMP “diagnosis” label does not correspond to clinical diagnosis, and is often incorrectly referred to as such given the nomenclature of the variable. Moreover, the KSADS-COMP-generated ED “diagnoses” are reported as “present” and “past”, where “present” indicates the past two weeks, and “past” reflects at any point in their lives (‘ever’).

### ED_SI_ nomenclature creation

The current manuscript uses the KSADS-COMP item-level data to develop new *DSM-5*-aligned ED_SI_ (e.g., Binge Eating Disorder Symptom Index [BED_SI_]). Given the nature of the KSADS-COMP item-level data, categorization of the ED_SI_ approximated the *DSM-5* inclusion criteria as closely as possible while allowing greater flexibility to capture the greatest number of youth as possible exhibiting potential ED symptoms. We refrained from using the terms “diagnosis”, “diagnostic,” and similar language when referring to the KSADS-COMP-derived categories or these new ED_SI_. We included the following ED_SI_ categories: anorexia nervosa restricting type (AN-R_SI_), anorexia nervosa binge/purge type (AN-BP_SI_), including atypical anorexia nervosa (AAN-R_SI_, AAN-BP_SI_) and anorexia nervosa-weight criteria unknown (AN-R-wcu_SI_, AN-BP-wcu_SI_) for both subtypes, bulimia nervosa (BN_SI_), and binge eating disorder (BED_SI_). OSFED-AN-R_SI_ and OSFED-BED_SI_ were generated for youth who did not meet the full criteria for AN-R_SI_ and BED_SI_, respectively, but still exhibited some symptoms (see below for how each category was determined). Due to KSADS-COMP question wording, it was impossible to distinguish between OSFED-AN-BP and OSFED-BN; therefore, they were combined into a single OSFED-AN-BP/BN_SI_ category. Additional youth who exhibited weight-control symptoms without endorsing specific weight-control methods were included in the OSFED-general_SI_ category. Notably, this manuscript focuses on reporting “present” symptomatology; we intentionally excluded questions that probed "past" symptoms, given ambiguity in the timeframe that those questions aimed to target. See **Figure 1** for a schematic of ED_SI_ categories and their KSADS-COMP counterparts described in this manuscript.

**Figure 1.**
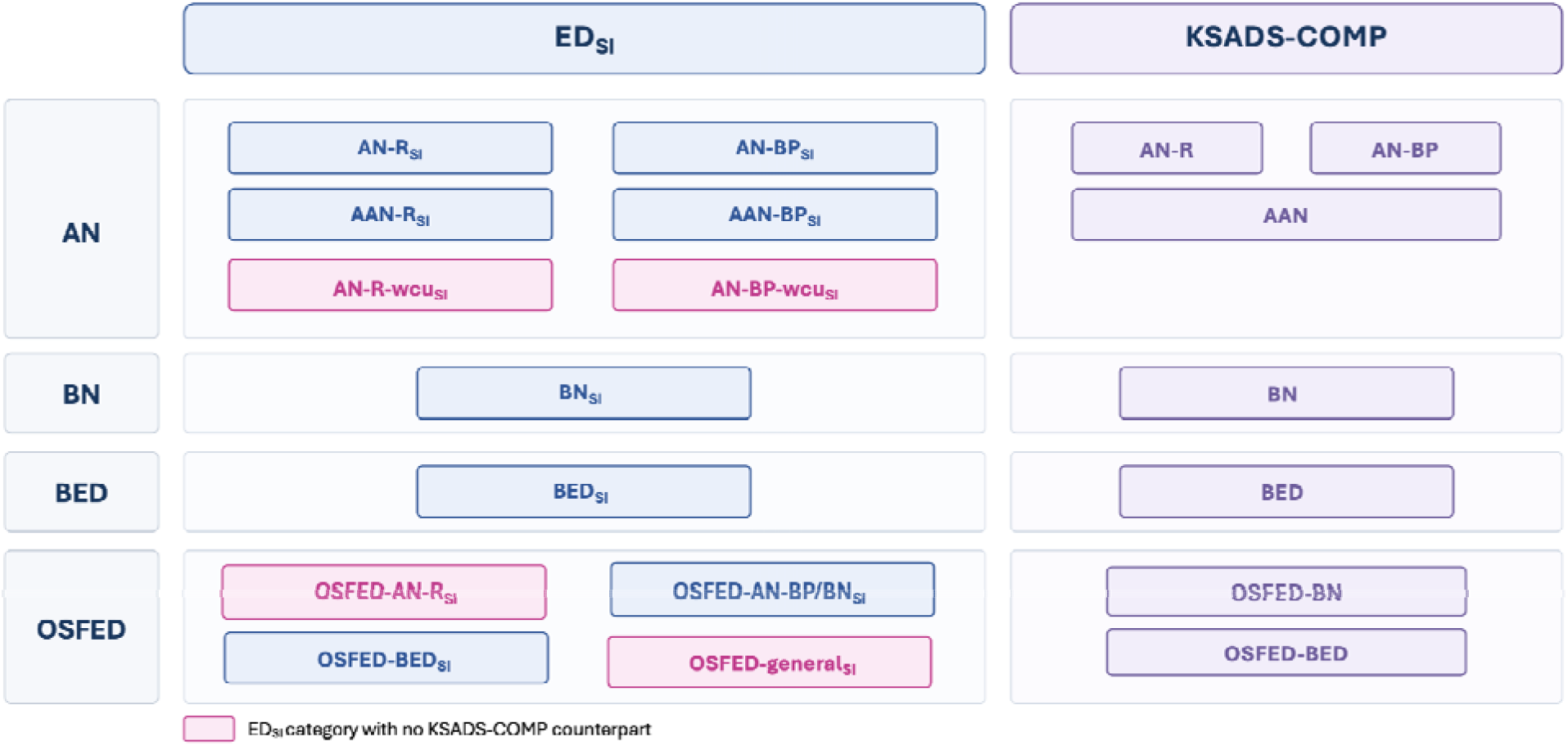
Schematic of ED_SI_ and KSADS-COMP ED categories. Abbreviations: ED, eating disorder; SI, Symptom Index; KSADS-COMP, Kiddie Schedule for Affective Disorders and Schizophrenia - Computerized Version; AN, Anorexia Nervosa; AAN, Atypical Anorexia Nervosa; R, Restricting Subtype; BP, Binge-purge Subtype; wcu, weight criterion unknown; BN, Bulimia Nervosa; BED, Binge Eating Disorder; OSFED, Other Specified Feeding and Eating Disorder.

### Anorexia Nervosa Symptom Index (AN_SI_)

*DSM-5* diagnostic criteria (American Psychiatric Association, 2022) for AN require the following criteria to be met: fear of weight gain (Criterion A), weight distress (Criterion B), significantly low body weight status (Criterion C), compensatory weight-control behaviors must have occurred at least once per week (Criterion D), and for at least 3 months (Criterion E). The corresponding item-level questions that best mapped onto these are outlined in **Table 1**. Subtype classifications were based on the presence or absence of binge-eating or purging behaviors (Abber et al., 2025, 2026; American Psychiatric Association, 2022). AN-R_SI_ criterion includes endorsement of at least one restrictive weight-control method (Criterion F.a) without any purging or binge eating (Criterion F.b). To meet the AN-BP_SI_ criteria, at least one purging weight-control method (Criterion G) must be reported.

**Table 1.**
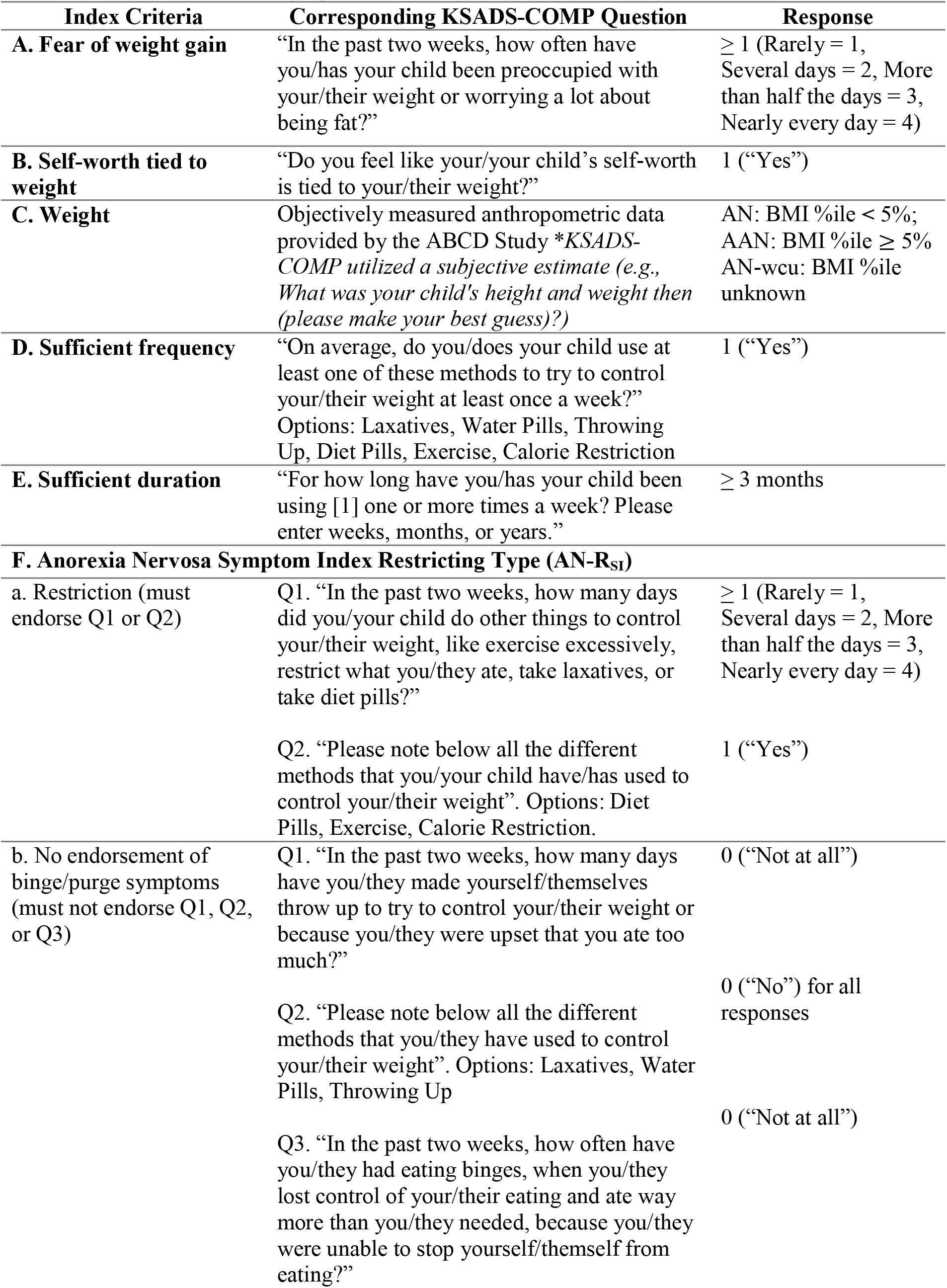
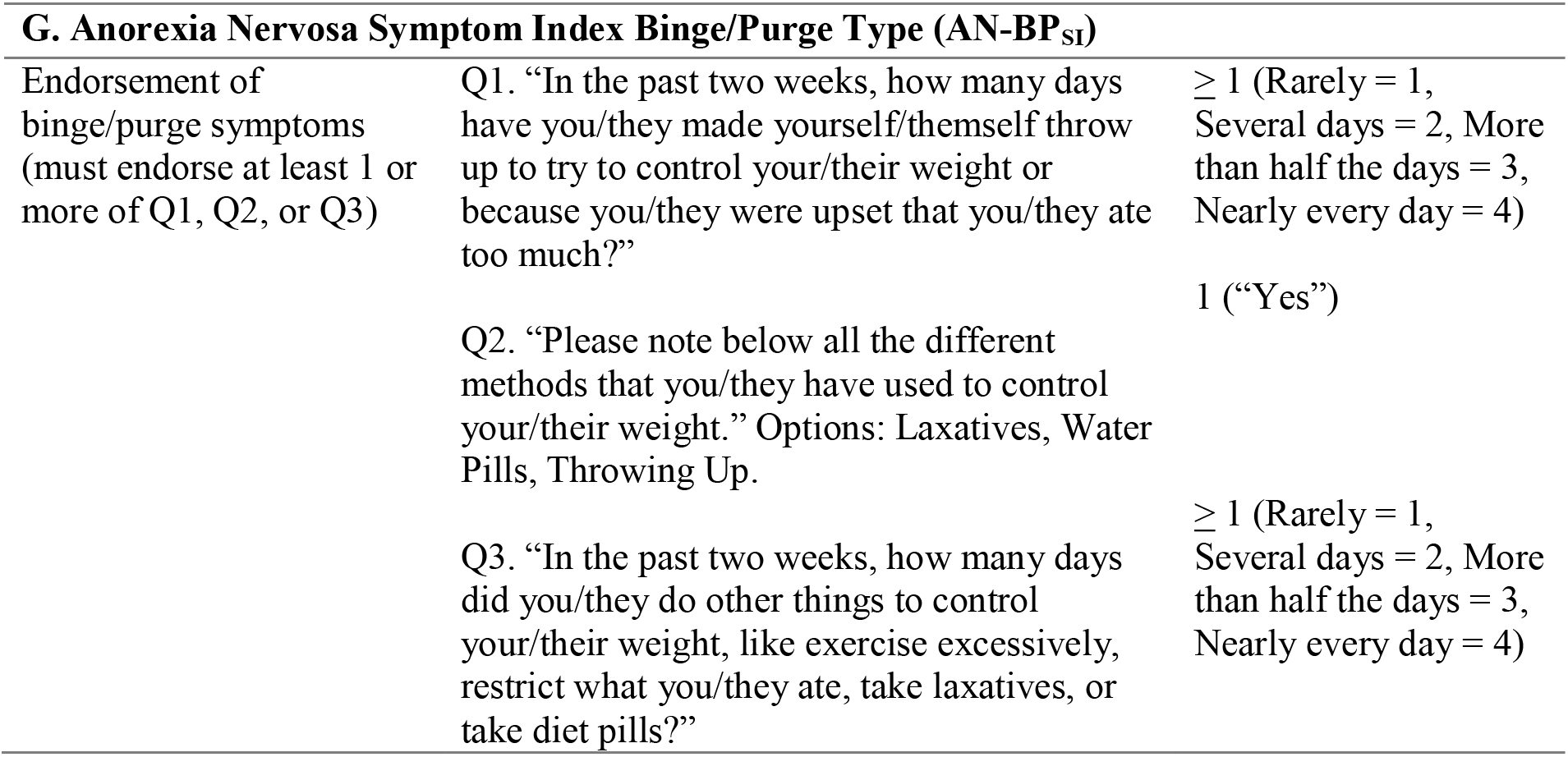
Anorexia Nervosa Symptom Index (AN_SI_). Questions asked in the KSADS-COMP youth-report ED module are specified. Caregivers completed the same questions, with the wording adjusted from “you” to “your child” or “they,” as appropriate. Each table outlines youth and caregiver language. “Q” refers to questions where the number reflects the sequence.

Notably, for our criteria, participants reporting both restrictive weight-control methods and binge/purge methods were categorized as AN-BP_SI_. As the KSADS-COMP combines both restrictive behaviors (e.g., limiting calorie intake) and purging behaviors (e.g., self-induced vomiting) within a single question (**Table 1**, Criterion F.a, Question 1), additional KSADS-COMP items (**Table 1**, Criterion F.b) assessing binge/purge behaviors (e.g., vomiting, laxatives, water pills, binge eating) were incorporated to distinguish endorsements of restrictive versus binge/purge behaviors.

We determined significantly low body weight (i.e., underweight) using objectively measured anthropometric data (i.e., height and weight) to calculate body mass index (BMI) percentiles based on the Centers for Disease Control’s sex- and age-specific growth charts (Kuczmarski et al., 2002). This was used to determine if a youth met AN (i.e., BMI < 5th percentile) or Atypical AN (AAN) (i.e., BMI typically in the normal weight range). A third AN_SI_ category was also created to capture youth where the weight criterion was unknown (AN-wcu_SI_); specifically, this captured youth meeting behavioral criteria for AN_SI_ but had either missing BMI data, biologically implausible values (e.g., measurement error), or only had subjectively reported BMI data (e.g., due to remote self-report assessments during the COVID-19 pandemic). We aimed to classify various AN_SI_ subgroups regardless of valid BMI data, given that ED-related psychopathology can be as severe among individuals with AAN as AN (Fitterman-Harris et al., 2024; Harrop et al., 2021; Johnson-Munguia, Bottera, et al., 2024; Johnson-Munguia, Negi, et al., 2024; Vo & Golden, 2022; Walsh et al., 2023).

### Bulimia Nervosa Symptom Index (BN_SI_)

*DSM-5* diagnostic criteria for bulimia nervosa (American Psychiatric Association, 2022) require recurrent binge eating episodes (Criterion A), compensatory weight control methods (Criterion B), weight distress (Criterion C), and sufficient frequency and duration of both binge-eating and weight control behaviors (Criteria D-G); **Table 2** details overlapping questions from the KSADS-COMP.

**Table 2.**
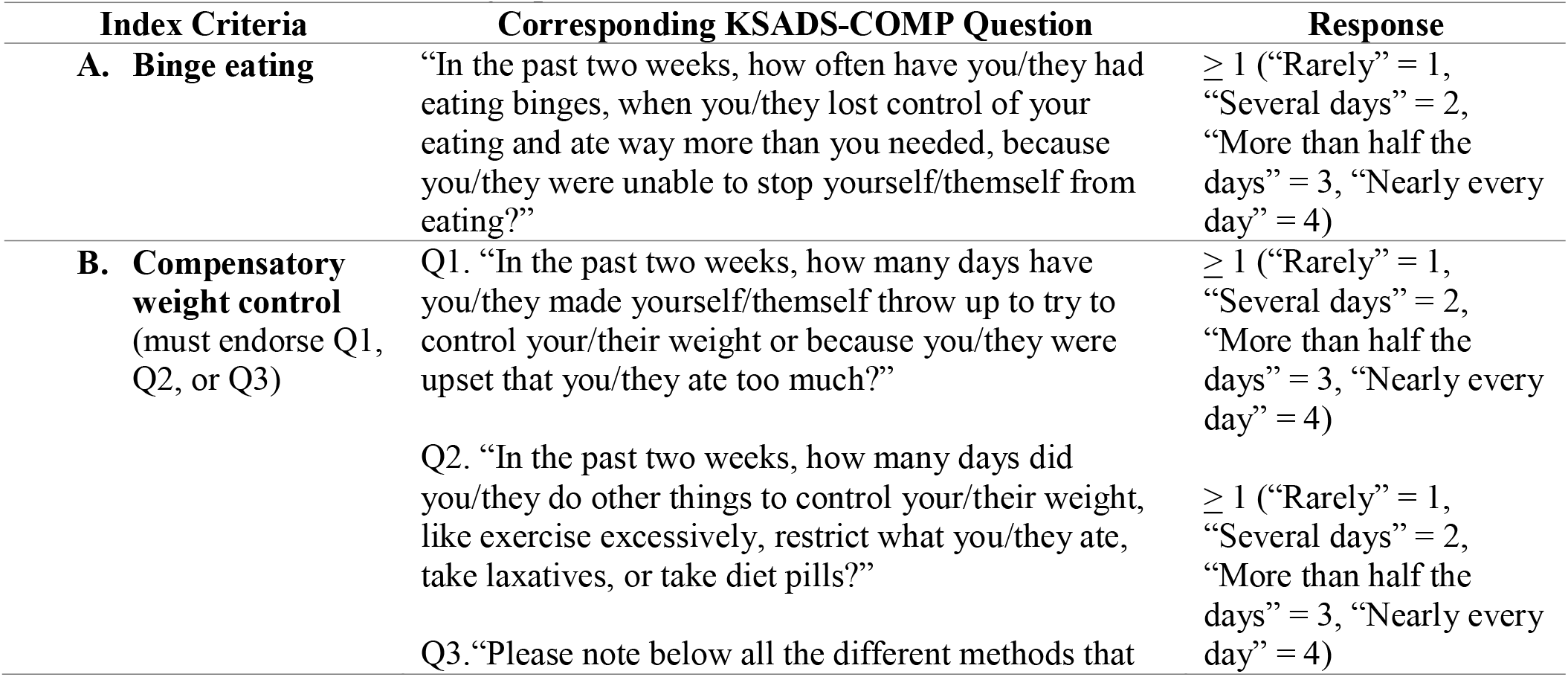
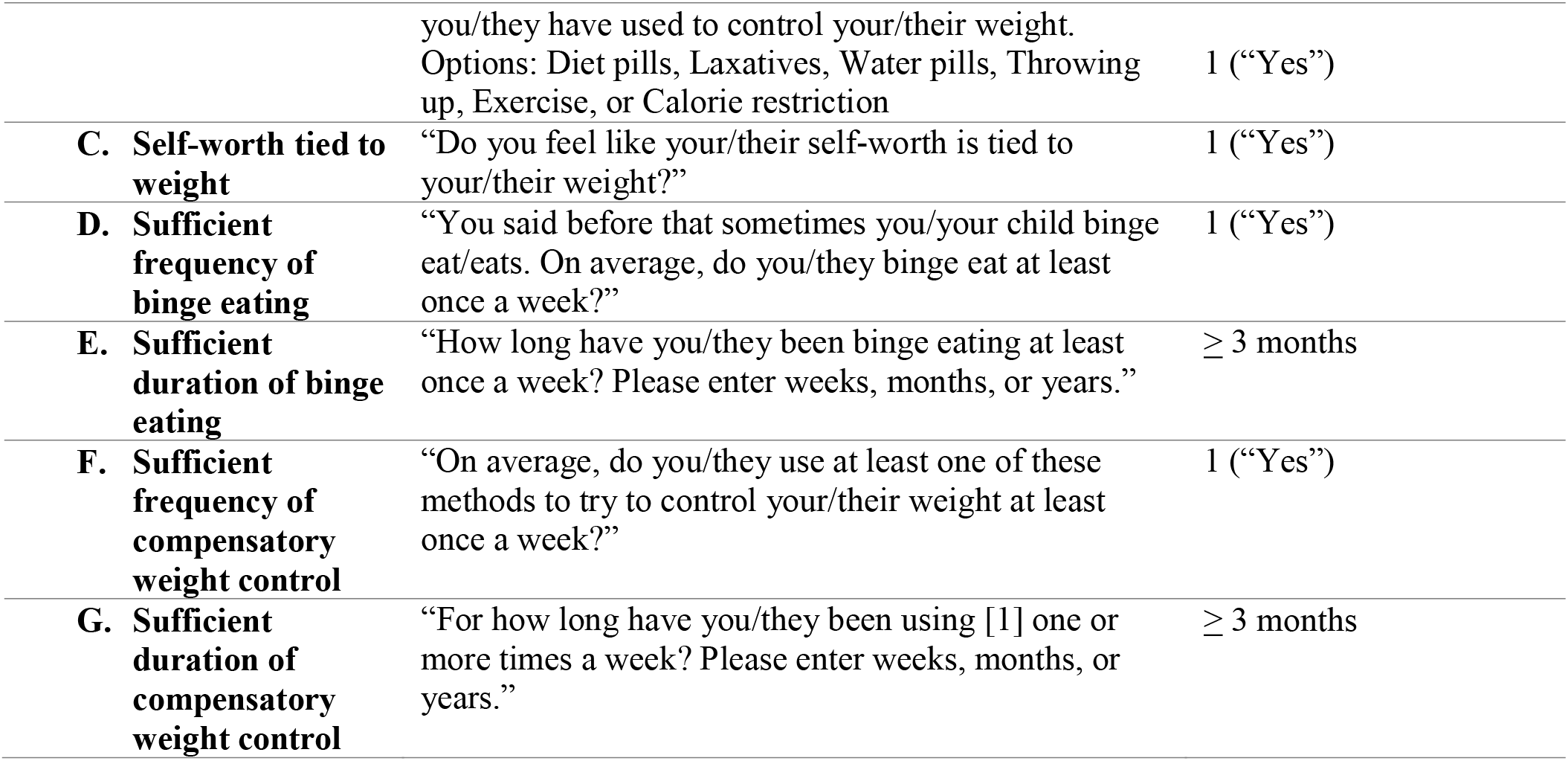
Bulimia Nervosa Symptom Index (BN_SI_). Questions asked in the KSADS-COMP youth-report ED module are specified. Caregivers completed the same questions, with the wording adjusted from “you” to “your child” or “they,” as appropriate. Each table outlines youth and caregiver language. “Q” refers to questions where the number reflects the sequence.

| Index Criteria | Corresponding KSADS-COMP Question | Response |
|---|---|---|
| **A. Binge eating** | “In the past two weeks, how often have you/they had eating binges, when you/they lost control of your eating and ate way more than you needed, because you/they were unable to stop yourself/themself from eating?” | ≥ 1 (“Rarely” = 1, “Several days” = 2, “More than half the days” = 3, “Nearly every day” = 4) |
| **B. Compensatory weight control** (must endorse Q1, Q2, or Q3) | Q1. “In the past two weeks, how many days have you/they made yourself/themself throw up to try to control your/their weight or because you/they were upset that you/they ate too much?” | ≥ 1 (“Rarely” = 1, “Several days” = 2, “More than half the days” = 3, “Nearly every day” = 4) |
|  | Q2. “In the past two weeks, how many days did you/they do other things to control your/their weight, like exercise excessively, restrict what you/they ate, take laxatives, or take diet pills?” | ≥ 1 (“Rarely” = 1, “Several days” = 2, “More than half the days” = 3, “Nearly every day” = 4) |
|  | Q3.“Please note below all the different methods that you/they have used to control your/their weight. Options: Diet pills, Laxatives, Water pills, Throwing up, Exercise, or Calorie restriction | 1 (“Yes”) |
| **C. Self-worth tied to weight** | “Do you feel like your/their self-worth is tied to your/their weight?” | 1 (“Yes”) |
| **D. Sufficient frequency of binge eating** | “You said before that sometimes you/your child binge eat/eats. On average, do you/they binge eat at least once a week?” | 1 (“Yes”) |
| **E. Sufficient duration of binge eating** | “How long have you/they been binge eating at least once a week? Please enter weeks, months, or years.” | ≥ 3 months |
| **F. Sufficient frequency of compensatory weight control** | “On average, do you/they use at least one of these methods to try to control your/their weight at least once a week?” | 1 (“Yes”) |
| **G. Sufficient duration of compensatory weight control** | “For how long have you/they been using [1] one or more times a week? Please enter weeks, months, or years.” | ≥ 3 months |

### Binge Eating Disorder Symptom Index (BED_SI_)

*DSM-5* diagnostic criteria for binge eating disorder (American Psychiatric Association, 2022) require recurrent binge eating episodes (Criterion A), at least three symptoms of binge eating (Criterion B), distress while binge eating (Criterion C), sufficient frequency (Criterion D) and duration of symptoms (Criterion E), and no simultaneous compensatory weight control methods (Criterion F); **Table 3** details overlapping questions from the KSADS-COMP.

**Table 3.** Binge Eating Disorder Symptom Index (BED_SI_). Questions asked in the KSADS-COMP youth-report ED module are specified. Caregivers completed the same questions, with the wording adjusted from “you” to “your child” or “they,” as appropriate. Each table outlines youth and caregiver language. “Q” refers to questions where the number reflects the sequence.

| Index Criteria | Corresponding KSADS-COMP Question | Response |
|---|---|---|
| A. **Binge eating** | “In the past two weeks, how often have you/they had eating binges, when you/they lost control of your eating and ate way more than you/they needed, because you/they were unable to stop yourself from eating?” | ≥ 1 (“Rarely” = 1, “Several days” = 2, “More than half the days” = 3, “Nearly every day” = 4) |
| B. **Symptoms of binge eating** (Must endorse at least 3 symptoms of options 1-7) | “Note below all the things that are true when you/they binge eat.”<br>1. I/they eat much more rapidly than normal<br>2. I/they eat until I am uncomfortable, and I feel too full<br>3. I/they eat a lot even though I/they am/are not hungry<br>4. I/they eat alone because I/they am/are embarrassed<br>5. I/they feel disgusted or guilty after binge eating<br>6. I/they feel depressed after binge eating<br>7. I/they feel like I/they have no control when I/they binge eat. | At least 3 positive responses (1s = “Yes”) |
| C. **Distress while binge eating** | “How much discomfort or distress does binge eating cause you?” | ≥ 1 (on a scale of 1-10) |
| D. **Sufficient frequency of binge eating** | “You/they said before that sometimes you/they binge eat. On average, do you/they binge eat at least once a week?” | 1 (“Yes”) |
| E. **Sufficient duration of binge eating** | “How long have you/they been binge eating at least once a week? Please enter weeks, months, or years.” | ≥ 3 months |
| F. **No compensatory weight-control** (must not endorse Q1, Q2, Q3 Q4, OR Q5) | Q1. “In the past two weeks, how many days have you/they made yourself/themself throw up to try to control your/their weight or because you/they were upset that you/they ate too much?”<br>Q2. “In the past two weeks, how many days did you/they do other things to control your/their weight, like exercise excessively, restrict what you/they ate, take laxatives, or take diet pills?”<br>Q3. “Please note below all the different methods that you/they have used to control your/their weight.”<br>Options: Diet pills, Laxatives, Water pills, Throwing up, Exercise, or Calorie restriction<br>Q4. “On average, do you/they use at least one of these methods to try to control your weight at least once a week?”<br>Q5. “For how long have you/they been using [1] one or more times a week? Please enter weeks, months, or years.” | 0 (“Not at all”)<br>0 (“Not at all”)<br>0 (“No”) or NA<br>0 (“No”) or NA<br>0 (“No”) or NA |

### Other Specified Feeding or Eating Disorder Symptom Index (OSFED_SI_)

Subthreshold ED presentations (e.g., not meeting all criteria listed in Tables 1-3 but still endorsing ED symptoms) are captured by the OSFED symptom index. Although this category is vague in the *DSM-5*, we aimed to capture as many nuances as possible in subthreshold symptom presentations, allowing researchers the flexibility to merge or separate groupings relevant to their research questions (Jenkins et al., 2021). Thus, we created four OSFED_SI_ presentations: OSFED-AN-R_SI_, OSFED-AN-BP/BN_SI,_ OSFED-BED_SI_, and OSFED-general_SI_. The OSFED-AN-R_SI_ and OSFED-BED_SI_ categories capture presentations of AN-R_SI_ and BED_SI_ symptoms, respectively, where not all initial criteria were met. The OSFED-AN-BP/BN_SI_ category describes common presentations of binge-eating or the binge/purge cycle, but frequency and/or duration thresholds were not met. It was impossible to disentangle OSFED-AN-BP_SI_ and OSFED-BN_SI_ cases using the available KSADS-COMP data; therefore, they were combined into a single category. OSFED-general_SI_ captures any clinically significant eating pathology for participants not already captured by a more stringent symptom index or specific to any particular weight control method. By comparison, KSADS-COMP defines two OSFED categories: OSFED-BN and OSFED-BED. Full operational criteria are detailed in the Supplementary Materials (Tables S1-S4).

### Statistical Analysis

Data were prepared using Python 3.13.5 and pandas 2.3.1 for data cleaning and analysis. All criteria-relevant data were coded into categorical variables with short descriptive variable names. For example, a score of 1 or higher on the “fear of weight gain” question was coded as “fear_obese”. Participant-reported values for the duration of a given ED symptom were converted to numeric month values for the duration criteria. These variables were then used to define ED_SI_.

We compared the number of participants meeting the KSADS-COMP and ED_SI_ criteria across time points, from the 3-year follow-up onward (3-, 4-, 6-, and 7-year follow-ups in the caregiver-report, and 4- and 6-year follow-ups in the youth report). Results are reported for the number of youth who met the KSADS-COMP and ED_SI_ criteria, separated by caregiver- vs. youth-report, by year, and sex. In cases where at least 1 but fewer than 10 youth met criteria, exact cell counts were suppressed in accordance with best practices for reporting small cell sizes (Seastrom, 2017). We used the Jaccard index to quantify the overlap between ED classifications (i.e., the number of participants who met *both* the KSADS-COMP and ED_SI_ criteria divided by the number who met *either* criterion). Values closer to 0 indicate no overlap, whereas values closer to 1 indicate complete overlap. When applicable (i.e., when there were sufficient youth for a meaningful comparison), chi-squared tests of proportions (conducted in *R* version 4.5.1) were used to assess the statistical significance of differences between proportions obtained from the KSADS-COMP and the ED_SI_. Yates’ continuity corrections were automatically applied to account for the chi-squared test’s tendency to overestimate significance in tables with small cell counts. These results are also stratified by sex to account for established differences in ED pathology by biological sex (Allen et al., 2013; Sanzari et al., 2023).

## RESULTS

In the sections that follow, we first quantify the number of youth that meet criteria for any given ED_SI_ category, followed by the number of youth that meet criteria for the equivalent ED-related category by KSADS-COMP. Note, that these method comparisons are for descriptive purposes only, given that it was not possible to pinpoint exactly where our ED_SI_ definitions differed from KSADS-COMP given the proprietary nature of its branching logic.

### AN

#### Caregiver-report

At all time points, fewer than 10 youth met the criteria for AN-R_SI_, and no youth met criteria for AN-BP_SI_. In comparison, 35 youth met KSADS-COMP criteria for AN-R, and 13 youth met KSADS-COMP criteria for AN-BP (see **Tables 4, 5**; **Figures 2A, 2C** display frequency over time). There was minimal overlap between the ED_SI_ and KSADS-COMP ED criteria in some categories, as participants who met the AN-R_SI_ criteria were *not* captured by the KSADS-COMP.

**Figure 2.**
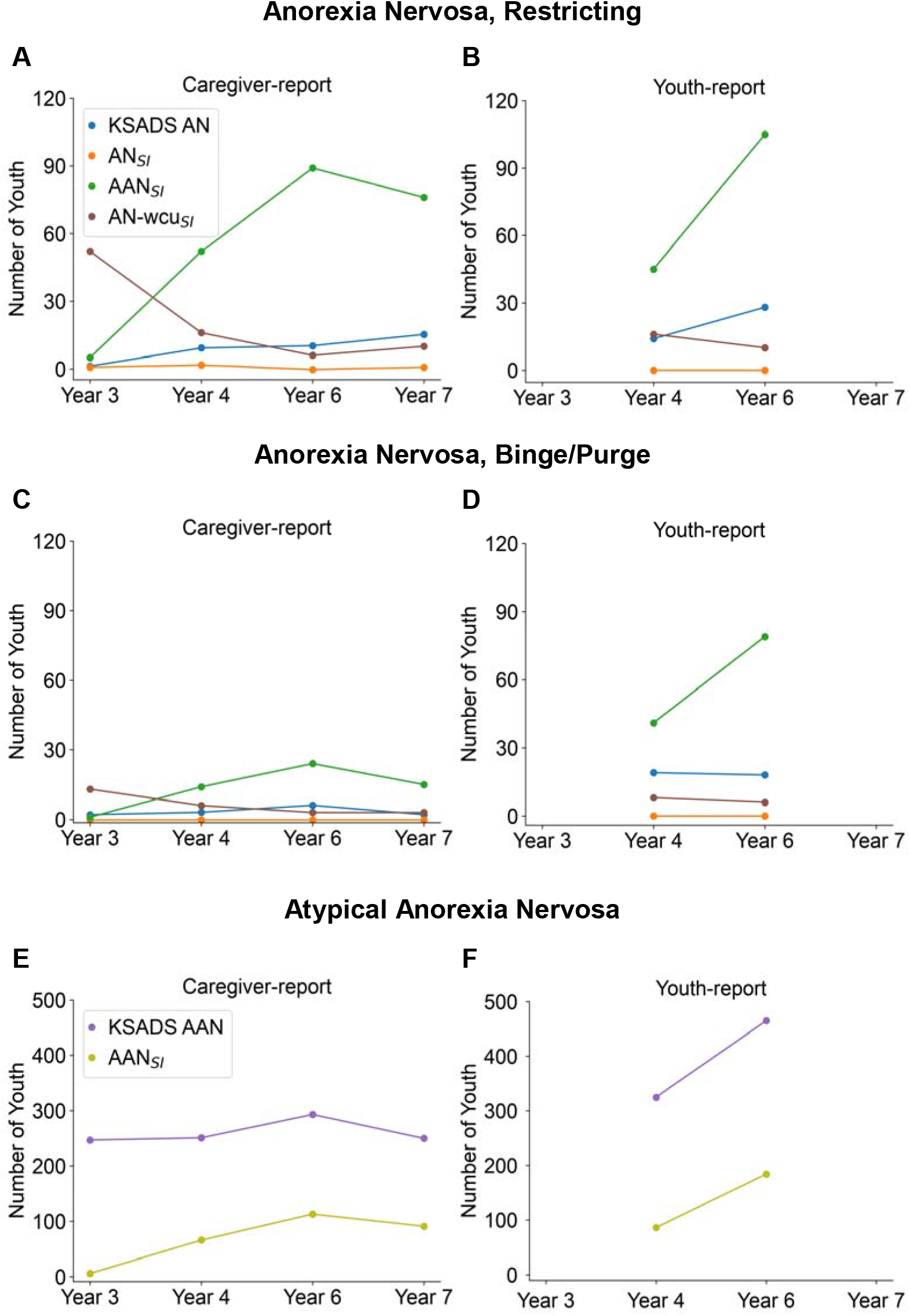
Number of youth who meet criteria for KSADS-COMP AN and AN_SI_. ***(A)*** Caregiver report of KSADS-COMP and ED_SI_ for AN-R. ***(B)*** Youth report of KSADS-COMP and ED_SI_ for AN-R. ***(C)*** Caregiver report of KSADS-COMP and ED_SI_ for AN-BP. ***(D)*** Youth report of KSADS-COMP and ED_SI_ for AN-BP. ***(E)*** Caregiver report of KSADS-COMP and ED_SI_ for AAN. ***(F)*** Youth report of KSADS-COMP and ED_SI_ for AAN.

**Table 4.**
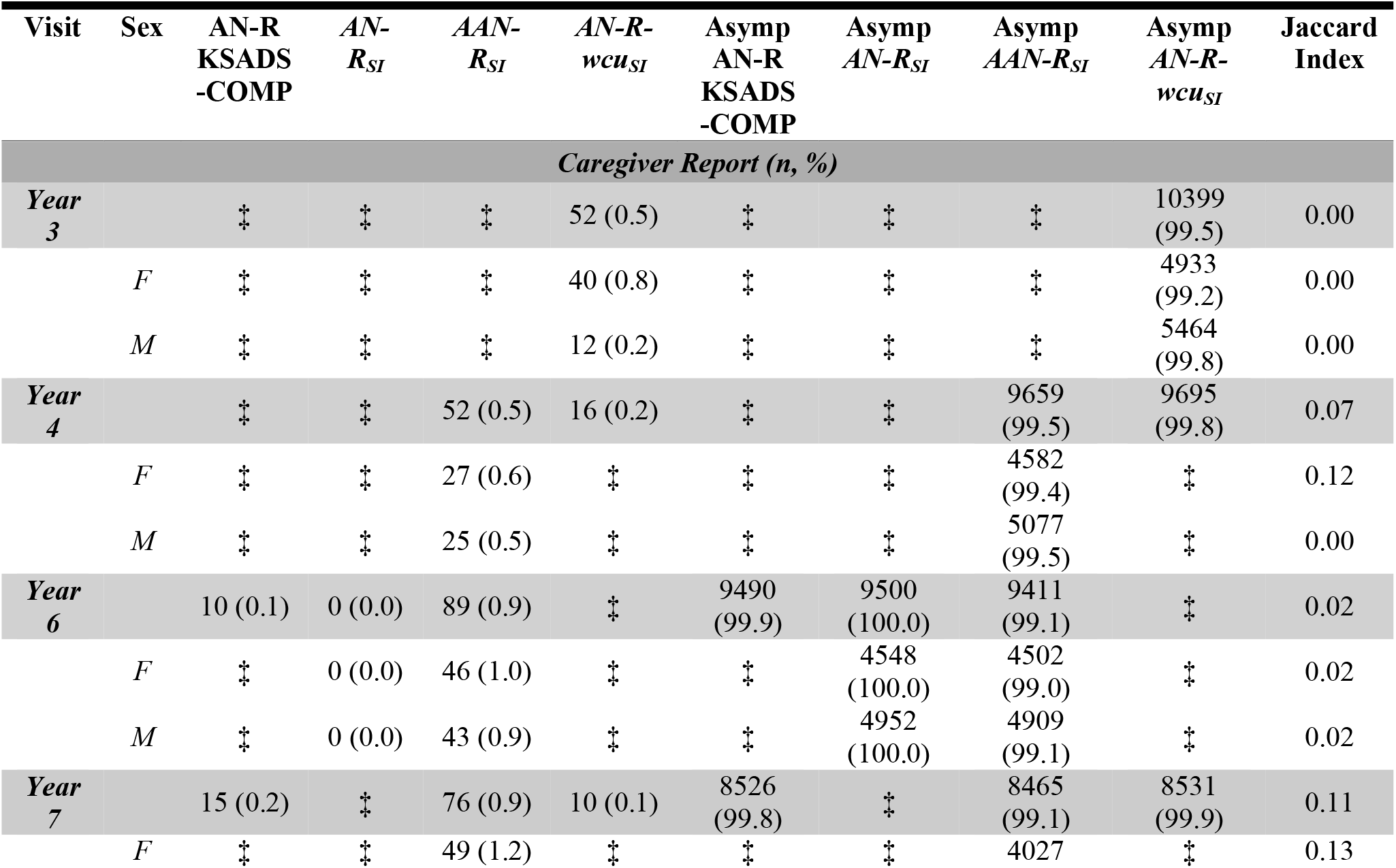
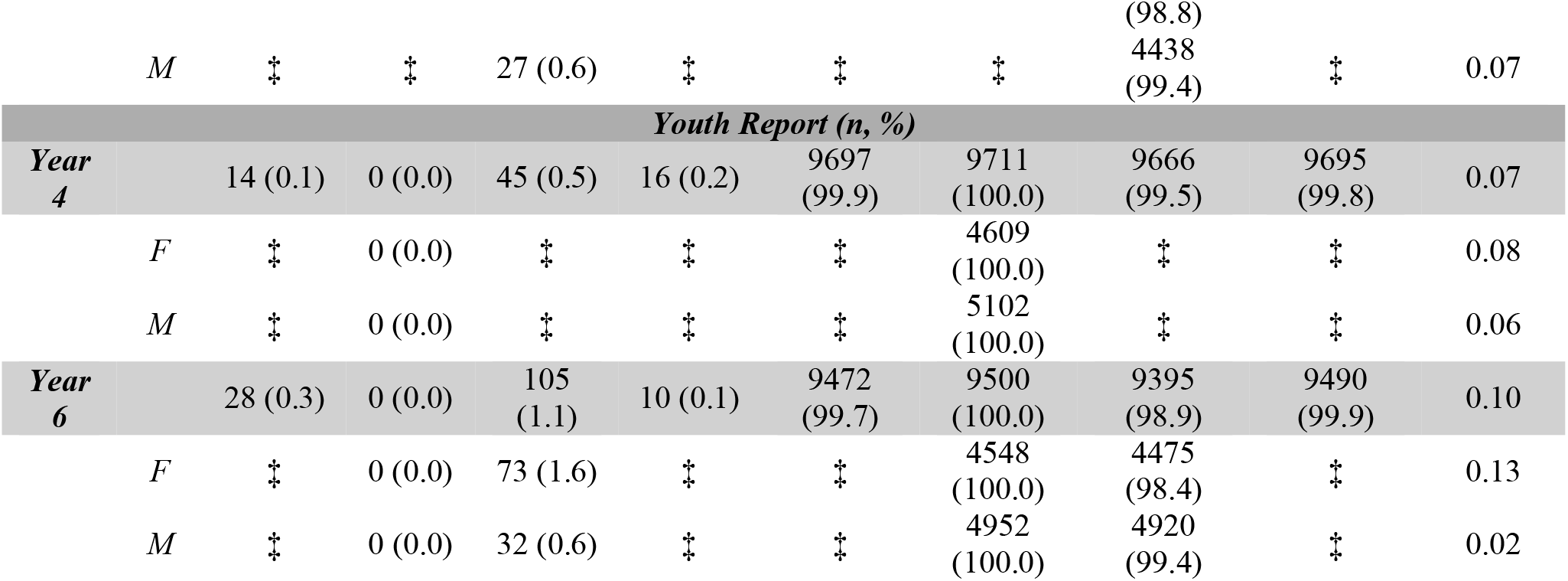
Number and proportion of youth that meet criteria for AN-R_SI_, AAN-R_SI_, and AN-R-wcu_SI_ compared to KSADS-COMP AN-R criteria. Data are presented as number of individuals meeting criteria and relative proportions in ABCD sample (%). AN-R_SI_ refers to youth who met criteria for the Anorexia Nervosa Symptom Index, restricting type. AAN-R_SI_ refers to youth who met criteria for the Atypical Anorexia Nervosa Symptom Index, restricting type. AN-R-wcu_SI_ refers to youth who met criteria for the Anorexia Nervosa Weight Criteria Unknown Symptom Index, restricting type. “Asymp” or “Asymptomatic” refers to youth who did not meet the criteria. Jaccard indices indicate the proportion of overlap between youth classified by both methods (KSADS-COMP vs. either AN-R_SI_, AAN-R_SI_, or AN-R-wcu_SI_). Chi-squared statistics were not included because the number of youth identified by the ED_SI_ classification system was too small for meaningful comparison. ‡ Reporting standards not met.

| **Visit** | **Sex** | **AN-R KSADS-COMP** | ***AN-R<sub>SI</sub>*** | ***AAN-R<sub>SI</sub>*** | ***AN-R-wcu<sub>SI</sub>*** | **Asymp AN-R KSADS-COMP** | **Asymp *AN-R<sub>SI</sub>*** | **Asymp *AAN-R<sub>SI</sub>*** | **Asymp *AN-R-wcu<sub>SI</sub>*** | **Jaccard Index** |
|---|---|---|---|---|---|---|---|---|---|---|
| ***Caregiver Report (n, %)*** |  |  |  |  |  |  |  |  |  |  |
| ***Year 3*** |  | ‡ | ‡ | ‡ | 52 (0.5) | ‡ | ‡ | ‡ | 10399 (99.5) | 0.00 |
|  | *F* | ‡ | ‡ | ‡ | 40 (0.8) | ‡ | ‡ | ‡ | 4933 (99.2) | 0.00 |
|  | *M* | ‡ | ‡ | ‡ | 12 (0.2) | ‡ | ‡ | ‡ | 5464 (99.8) | 0.00 |
| ***Year 4*** |  | ‡ | ‡ | 52 (0.5) | 16 (0.2) | ‡ | ‡ | 9659 (99.5) | 9695 (99.8) | 0.07 |
|  | *F* | ‡ | ‡ | 27 (0.6) | ‡ | ‡ | ‡ | 4582 (99.4) | ‡ | 0.12 |
|  | *M* | ‡ | ‡ | 25 (0.5) | ‡ | ‡ | ‡ | 5077 (99.5) | ‡ | 0.00 |
| ***Year 6*** |  | 10 (0.1) | 0 (0.0) | 89 (0.9) | ‡ | 9490 (99.9) | 9500 (100.0) | 9411 (99.1) | ‡ | 0.02 |
|  | *F* | ‡ | 0 (0.0) | 46 (1.0) | ‡ | ‡ | 4548 (100.0) | 4502 (99.0) | ‡ | 0.02 |
|  | *M* | ‡ | 0 (0.0) | 43 (0.9) | ‡ | ‡ | 4952 (100.0) | 4909 (99.1) | ‡ | 0.02 |
| ***Year 7*** |  | 15 (0.2) | ‡ | 76 (0.9) | 10 (0.1) | 8526 (99.8) | ‡ | 8465 (99.1) | 8531 (99.9) | 0.11 |
|  | *F* | ‡ | ‡ | 49 (1.2) | ‡ | ‡ | ‡ | 4027 (98.8) | ‡ | 0.13 |
|  | *M* | ‡ | ‡ | 27 (0.6) | ‡ | ‡ | ‡ | 4438 (99.4) | ‡ | 0.07 |
| ***Youth Report (n, %)*** |  |  |  |  |  |  |  |  |  |  |
| ***Year 4*** |  | 14 (0.1) | 0 (0.0) | 45 (0.5) | 16 (0.2) | 9697 (99.9) | 9711 (100.0) | 9666 (99.5) | 9695 (99.8) | 0.07 |
|  | *F* | ‡ | 0 (0.0) | ‡ | ‡ | ‡ | 4609 (100.0) | ‡ | ‡ | 0.08 |
|  | *M* | ‡ | 0 (0.0) | ‡ | ‡ | ‡ | 5102 (100.0) | ‡ | ‡ | 0.06 |
| ***Year 6*** |  | 28 (0.3) | 0 (0.0) | 105 (1.1) | 10 (0.1) | 9472 (99.7) | 9500 (100.0) | 9395 (98.9) | 9490 (99.9) | 0.10 |
|  | *F* | ‡ | 0 (0.0) | 73 (1.6) | ‡ | ‡ | 4548 (100.0) | 4475 (98.4) | ‡ | 0.13 |
|  | *M* | ‡ | 0 (0.0) | 32 (0.6) | ‡ | ‡ | 4952 (100.0) | 4920 (99.4) | ‡ | 0.02 |

**Table 5.** Number and proportion of youth that meet criteria for AN-BP_SI_ and AAN-BP_SI_ compared to KSADS-COMP AN-R criteria. Data are presented as number of individuals meeting criteria and relative proportions in ABCD sample (%). AN-BP_SI_ refers to youth who met criteria for the Anorexia Nervosa Symptom Index, binge/purge type. AAN-BP_SI_ refers to youth who met criteria for the Atypical Anorexia Nervosa Symptom Index, binge/purge type. AN-BP-wcu_SI_ refers to youth who met criteria for the Anorexia Nervosa Weight Criteria Unknown Symptom Index, binge/purge type. “Asymp” or “Asymptomatic” refers to youth who did not meet the criteria. Jaccard indices indicate the proportion of overlap between youth classified by both methods (KSADS-COMP vs. either AN-BP_SI_, AAN-BP_SI_, or AN-BP-wcu_SI_). Chi-squared statistics were not included because the number of youth identified by the ED_SI_ classification system was too small for meaningful comparison. ‡ Reporting standards not met.

| Visit | Sex | AN-BP KSADS -COMP | *AN-BP*<sub>SI</sub> | *AAN-BP*<sub>SI</sub> | *AN-BP-wcu<sub>SI</sub>* | Asymp AN-BP KSADS -COMP | Asymp *AN-BP<sub>SI</sub>* | Asymp *AAN-BP<sub>SI</sub>* | Asymp *AN-BP-wcu<sub>SI</sub>* | Jaccard Index |
|---|---|---|---|---|---|---|---|---|---|---|
| ***Caregiver Report (n, %)*** |  |  |  |  |  |  |  |  |  |  |
| ***Year 3*** |  | ‡ | 0 (0.0) | ‡ | 13 (0.1) | ‡ | 10451 (100.0) | ‡ | 10438 (99.9) | 0.00 |
|  | *F* | ‡ | 0 (0.0) | ‡ | ‡ | ‡ | 4973 (100.0) | ‡ | ‡ | 0.00 |
|  | *M* | ‡ | 0 (0.0) | ‡ | ‡ | ‡ | 5476 (100.0) | ‡ | ‡ | 0.00 |
| ***Year 4*** |  | ‡ | 0 (0.0) | 14 (0.1) | ‡ | ‡ | 9711 (100.0) | 9697 (99.9) | ‡ | 0.05 |
|  | *F* | ‡ | 0 (0.0) | ‡ | ‡ | ‡ | 4609 (100.0) | ‡ | ‡ | 0.07 |
|  | *M* | ‡ | 0 (0.0) | ‡ | ‡ | ‡ | 5102 (100.0) | ‡ | ‡ | 0.00 |
| ***Year 6*** |  | ‡ | 0 (0.0) | 24 (0.3) | ‡ | ‡ | 9500 (100.0) | 9476 (99.7) | ‡ | 0.18 |
|  | *F* | ‡ | 0 (0.0) | ‡ | ‡ | ‡ | 4548 (100.0) | ‡ | ‡ | 0.26 |
|  | *M* | ‡ | 0 (0.0) | ‡ | ‡ | ‡ | 4952 (100.0) | ‡ | ‡ | 0.00 |
| ***Year 7*** |  | ‡ | 0 (0.0) | 15 (0.2) | ‡ | ‡ | 8541 (100.0) | 8526 (99.8) | ‡ | 0.05 |
|  | *F* | ‡ | 0 (0.0) | ‡ | ‡ | ‡ | 4076 (100.0) | ‡ | ‡ | 0.07 |
|  | *M* | ‡ | 0 (0.0) | ‡ | ‡ | ‡ | 4465 (100.0) | ‡ | ‡ | 0.00 |
| ***Youth Report (n, %)*** |  |  |  |  |  |  |  |  |  |  |
| ***Year 4*** |  | 19 (0.2) | 0 (0.0) | 41 (0.4) | ‡ | 9692 (99.8) | 9711 (100.0) | 9670 (99.6) | ‡ | 0.10 |
|  | *F* | ‡ | 0 (0.0) | 30 (0.7) | ‡ | ‡ | 4609 (100.0) | 4579 (99.3) | ‡ | 0.13 |
|  | *M* | ‡ | 0 (0.0) | 11 (0.2) | ‡ | ‡ | 5102 (100.0) | 5091 (99.8) | ‡ | 0.00 |
| ***Year 6*** |  | 18 (0.2) | 0 (0.0) | 79 (0.8) | ‡ | 9482 (99.8) | 9500 (100.0) | 9421 (99.2) | ‡ | 0.06 |
|  | *F* | ‡ | 0 (0.0) | 62 (1.4) | ‡ | ‡ | 4548 (100.0) | 4486 (98.6) | ‡ | 0.06 |
|  | *M* | ‡ | 0 (0.0) | 17 (0.3) | ‡ | ‡ | 4952 (100.0) | 4935 (99.7) | ‡ | 0.05 |

The impact of objectively measured BMI vs. caregiver-reported estimates of height and weight on categorizations was tested. Jaccard indices were calculated between participants who met the KSADS-COMP criteria for AN-R (using self-reported anthropometrics) and those who met the ED_SI_ criteria for *either* AN-R_SI_, AAN-R_SI_, or AN-R-wcu_SI_ (using objective BMI data when possible) (see **Table 4**; see **Table 5** for the same analyses conducted across the binge/purge subtype). Jaccard indices ranged from 0.00 to 0.11 for the restricting subtype, and from 0.00 to 0.18 for the binge/purge subtype, indicating minimal overlap. Thus, differences in BMI calculations between the ED_SI_ and KSADS-COMP were not the primary cause of differences in categorization. However, the specific criteria and decision rules used in the KSADS-COMP to determine AN in this dataset were not transparent, limiting the ability to make a precise comparison.

#### Youth-report

Across all time points, no youth met the criteria for either AN-R_SI_ or AN-BP_SI_ (see **Tables 4, 5**; **Figures 2B, 2D** display frequency over time). Meanwhile, 42 youth met the KSADS-COMP criteria for AN-R, and 37 youth met the KSADS-COMP criteria for AN-BP. Similar comparative analyses were conducted using caregiver-report data, and the Jaccard indices ranged from 0.07 to 0.10 for the restricting subtype and from 0.06 to 0.10 for the binge-purge subtype. The minimal overlap similarly reveals that usage of objectively measured BMI accounted for only a few differences in classification.

### AAN

#### Caregiver-report

More youth met criteria for the AAN_SI_ than the AN_SI_. Across all time points, 222 youth met the criteria for AAN-R_SI_, and 54 met the criteria for AAN-BP_SI_ (see **Tables 4 and 5**; **Figures 2A and 2C** show frequencies over time). Direct comparison with the KSADS-COMP by subtype was not possible because the KSADS-COMP reports only a broad AAN category rather than separate AAN-R and AAN-BP classifications. Accordingly, to conduct comparative analyses, we combined youth who met criteria for either AAN-R_SI_ or AAN-BP_SI_ and compared this aggregated classification with the KSADS-COMP AAN classification (see **Table 6**; **Figure 2E** displays frequency over time). Results indicated minimal overlap between youth identified by AAN_SI_ criteria and those meeting KSADS-COMP criteria for AAN, with Jaccard indices ranging from 0.02 to 0.35.

**Table 6.**
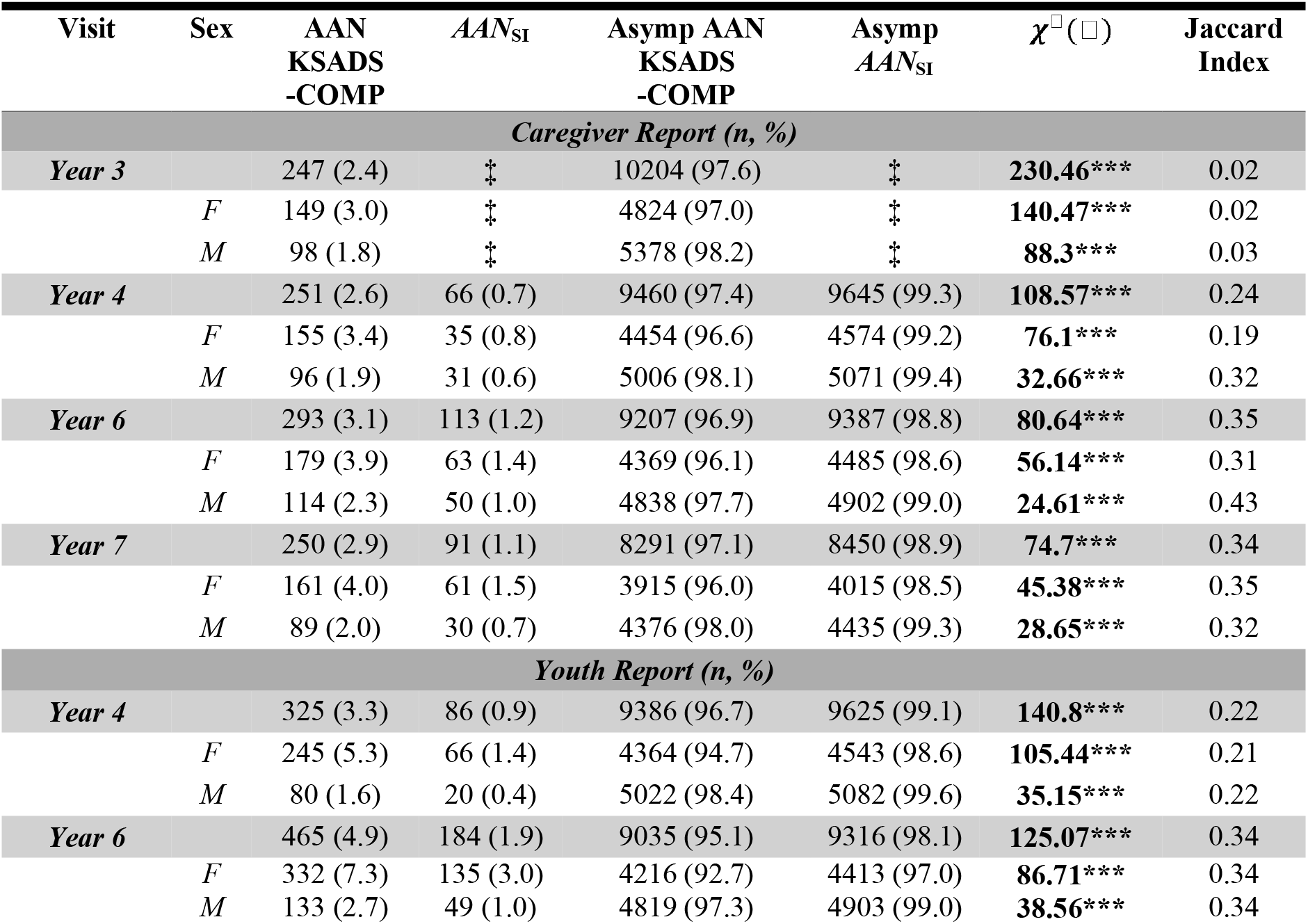
Number and proportion of youth who met criteria for AAN_SI_ (summed across subtypes) compared to KSADS-COMP AAN criteria. Data are presented as number of individuals meeting criteria and relative proportions in ABCD sample (%). AAN_SI_ refers to youth who met the criteria for the Atypical Anorexia Nervosa Symptom Index, either subtype. “Asymp” or “Asymptomatic” refers to youth who did not meet the criteria. Jaccard indices indicate the proportion of overlap between youth classified by both methods (KSADS-COMP vs. AAN_SI_). Chi-squared statistics reflect tests of proportions for the KSADS-COMP vs. AAN_SI_ at each time point, stratified by sex, with 1 degree of freedom and Yates continuity correction applied where possible. F = female; M = male; *** = p < 0.001; ** = p < 0.01; * = p < 0.05.‡ Reporting standards not met.

| Visit | Sex | AAN<br>KSADS<br>-COMP | *AAN*<sub>SI</sub> | Asymp AAN<br>KSADS<br>-COMP | Asymp<br>*AAN*<sub>SI</sub> | *χ*<sup>□</sup>(□) | Jaccard<br>Index |
|---|---|---|---|---|---|---|---|
| ***Caregiver Report (n, %)*** |  |  |  |  |  |  |  |
| ***Year 3*** |  | 247 (2.4) | ‡ | 10204 (97.6) | ‡ | **230.46\*\*\*** | 0.02 |
|  | *F* | 149 (3.0) | ‡ | 4824 (97.0) | ‡ | **140.47\*\*\*** | 0.02 |
|  | *M* | 98 (1.8) | ‡ | 5378 (98.2) | ‡ | **88.3\*\*\*** | 0.03 |
| ***Year 4*** |  | 251 (2.6) | 66 (0.7) | 9460 (97.4) | 9645 (99.3) | **108.57\*\*\*** | 0.24 |
|  | *F* | 155 (3.4) | 35 (0.8) | 4454 (96.6) | 4574 (99.2) | **76.1\*\*\*** | 0.19 |
|  | *M* | 96 (1.9) | 31 (0.6) | 5006 (98.1) | 5071 (99.4) | **32.66\*\*\*** | 0.32 |
| ***Year 6*** |  | 293 (3.1) | 113 (1.2) | 9207 (96.9) | 9387 (98.8) | **80.64\*\*\*** | 0.35 |
|  | *F* | 179 (3.9) | 63 (1.4) | 4369 (96.1) | 4485 (98.6) | **56.14\*\*\*** | 0.31 |
|  | *M* | 114 (2.3) | 50 (1.0) | 4838 (97.7) | 4902 (99.0) | **24.61\*\*\*** | 0.43 |
| ***Year 7*** |  | 250 (2.9) | 91 (1.1) | 8291 (97.1) | 8450 (98.9) | **74.7\*\*\*** | 0.34 |
|  | *F* | 161 (4.0) | 61 (1.5) | 3915 (96.0) | 4015 (98.5) | **45.38\*\*\*** | 0.35 |
|  | *M* | 89 (2.0) | 30 (0.7) | 4376 (98.0) | 4435 (99.3) | **28.65\*\*\*** | 0.32 |
| ***Youth Report (n, %)*** |  |  |  |  |  |  |  |
| ***Year 4*** |  | 325 (3.3) | 86 (0.9) | 9386 (96.7) | 9625 (99.1) | **140.8\*\*\*** | 0.22 |
|  | *F* | 245 (5.3) | 66 (1.4) | 4364 (94.7) | 4543 (98.6) | **105.44\*\*\*** | 0.21 |
|  | *M* | 80 (1.6) | 20 (0.4) | 5022 (98.4) | 5082 (99.6) | **35.15\*\*\*** | 0.22 |
| ***Year 6*** |  | 465 (4.9) | 184 (1.9) | 9035 (95.1) | 9316 (98.1) | **125.07\*\*\*** | 0.34 |
|  | *F* | 332 (7.3) | 135 (3.0) | 4216 (92.7) | 4413 (97.0) | **86.71\*\*\*** | 0.34 |
|  | *M* | 133 (2.7) | 49 (1.0) | 4819 (97.3) | 4903 (99.0) | **38.56\*\*\*** | 0.34 |

#### Youth-report

The frequency of AAN-R_SI_ and AAN-BP_SI_ was also higher than that of AN-R_SI_ and AN-BP_SI_, respectively, with 150 youth meeting criteria for AAN-R_SI_ and 120 for AAN-BP_SI_ (see **Tables 4, 5**; **Figures 2B, 2D** display frequency over time). As with caregiver-report analyses, direct comparison with the KSADS-COMP was not possible because it reports only an AAN category. Therefore, we conducted analogous analyses using the broader AAN classification reported by the KSADS-COMP (see **Table 6**; **Figure 2F** displays frequency over time). Similar to caregiver-report findings, there was minimal overlap between youth identified by the two classification approaches, with Jaccard indices ranging from 0.22 to 0.34.

### AN-wcu

#### Caregiver-report

Across all time points, 84 youth met criteria for AN-R-wcu_SI_, and 25 youth met criteria for AN-BP-wcu_SI_ (see **Tables 4, 5**; **Figures 2A, 2C** display frequency over time). The number of participants who met criteria for AN-BP-wcu_SI_ and AN-R-wcu_SI_ was consistently highest at the 3-year follow-up. This could be attributable to the fact that most participants completed their assessments remotely due to COVID-19. Thus, their objective BMI data were not collected, resulting in categorization of AN-wcu_SI_.

#### Youth-report

Across all time points, 26 youth met criteria for AN-R-wcu_SI_, and 14 youth met criteria for AN-BP-wcu_SI_ (see **Tables 4, 5**; **Figures 2B, 2D** display frequency over time).

Because youth did not complete a self-report at the 3-year follow-up, these data were not impacted by COVID-19-related constraints. Therefore, we do not expect this number to be higher, as was observed for caregiver reports.

### BN

#### Caregiver-report

Across all time points, 65 youth met the BN_SI_ criteria, whereas 60 youth met the KSADS-COMP criteria for BN. Jaccard indices ranged from 0.88 to 0.95, indicating substantial agreement between the BN_SI_ results and the KSADS-COMP-derived BN classifications (see **Table 7**; **Figure 3A** displays frequency over time). All youth in the BN_SI_ criteria were identified in the KSADS-COMP criteria, however, additional youth were also captured in the BN_SI_ criteria.

**Table 7.** Number and proportion of youth who met criteria for BN_SI_ compa red to KSAD S-COMP BN criteria. Data are presented as number of individuals meeting criteria and relative proportions in ABCD sample (%). BN_SI_ reflects youth who met the criteria for the Bulimia Nervosa Symptom Index. “Asymp” or “Asymptomatic” reflects youth who did not meet the criteria. Jaccard indices indicate the proportion of overlap in youth classified by both methods (KSADS-COMP vs. BN_SI_). Chi-squared statistics reflect tests of proportions for the KSADS-COMP vs. BN_SI_ at each time point, stratified by sex, with 1 degree of freedom and Yates continuity correction applied where possible. F = female; M = male; *** = p < 0.001; ** = p < 0.01; * = p < 0.05. ‡ Reporting standards not met

| Visit | Sex | **BN KSADS-COMP** | ***BN*<sub>SI</sub>** | **Asymp BN KSADS-COMP** | **Asymp *BN*<sub>SI</sub>** | χ<sup>2</sup>(1) | **Jaccard Index** |
|---|---|---|---|---|---|---|---|
| ***Caregiver Report (n, %)*** |  |  |  |  |  |  |  |
| ***Year 3*** |  | ‡ | ‡ | ‡ | ‡ | 0 | 0.89 |
|  | *F* | ‡ | ‡ | ‡ | ‡ | 0 | 1.00 |
|  | *M* | ‡ | ‡ | ‡ | ‡ | 0 | 0.75 |
| ***Year 4*** |  | 15 (0.20) | 17 (0.20) | 9696 (99.8) | 9694 (99.8) | 0.03 | 0.88 |
|  | *F* | ‡ | ‡ | ‡ | ‡ | 0 | 0.89 |
|  | *M* | ‡ | ‡ | ‡ | ‡ | 0 | 0.88 |
| ***Year 6*** |  | 18 (0.20) | 19 (0.20) | 9482 (99.8) | 9481 (99.8) | 0 | 0.95 |
|  | *F* | ‡ | ‡ | ‡ | ‡ | 0 | 1.00 |
|  | *M* | ‡ | ‡ | ‡ | ‡ | 0 | 0.83 |
| ***Year 7*** |  | 19 (0.20) | 20 (0.20) | 8522 (99.8) | 8521 (99.8) | 0 | 0.95 |
|  | *F* | ‡ | ‡ | ‡ | ‡ | 0 | 0.92 |
|  | *M* | ‡ | ‡ | ‡ | ‡ | 0 | 1.00 |
| ***Youth Report (n, %)*** |  |  |  |  |  |  |  |
| ***Year 4*** |  | 28 (0.30) | 32 (0.30) | 9683 (99.7) | 9679 (99.7) | 0.15 | 0.88 |
|  | *F* | ‡ | ‡ | ‡ | ‡ | 0.09 | 0.88 |
|  | *M* | ‡ | ‡ | ‡ | ‡ | 0 | 0.86 |
| ***Year 6*** |  | 62 (0.70) | 68 (0.70) | 9438 (99.3) | 9432 (99.3) | 0.19 | 0.91 |
|  | *F* | 49 (1.10) | 53 (1.20) | 4499 (98.9) | 4495 (98.8) | 0.09 | 0.92 |
|  | *M* | 13 (0.30) | 15 (0.30) | 4939 (99.7) | 4937 (99.7) | 0.04 | 0.87 |

‡ Reporting standards not met

**Figure 3.**
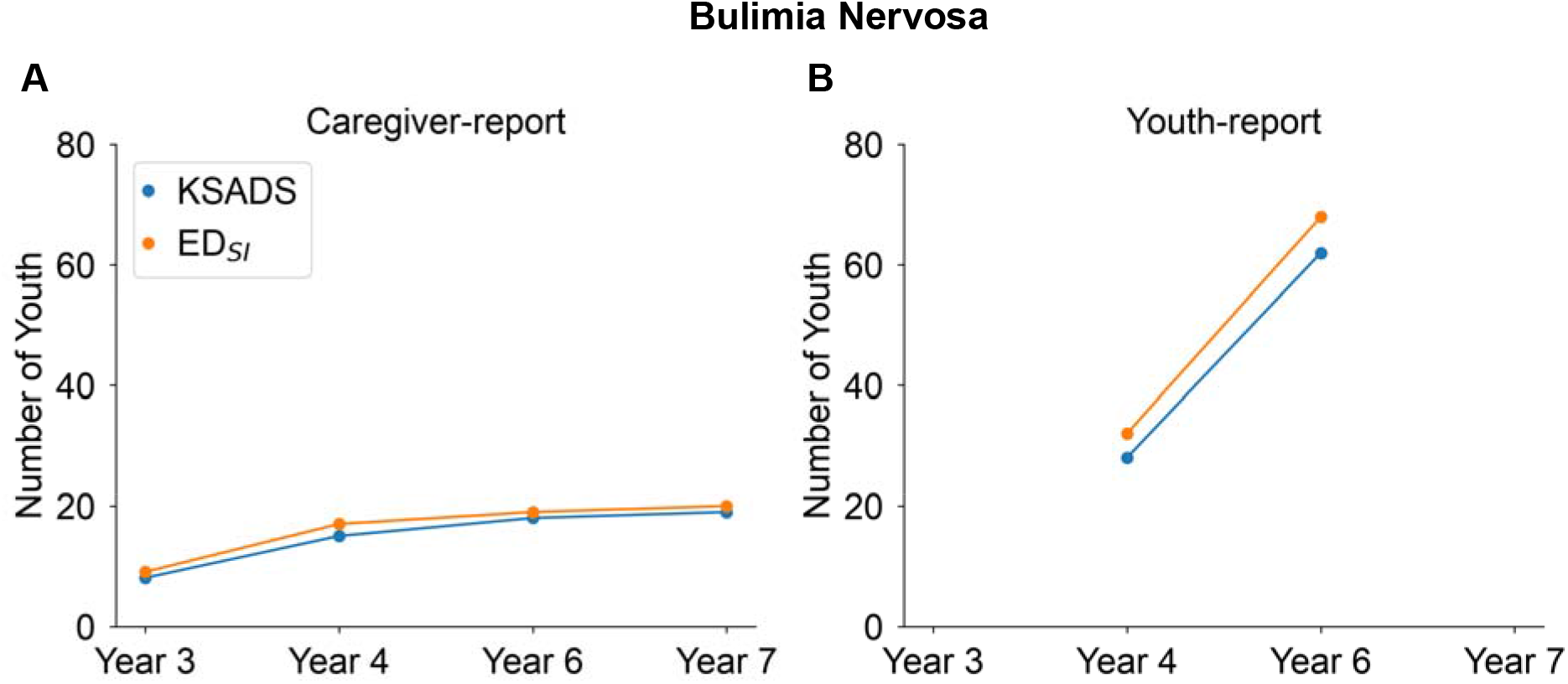
Number of youth who met criteria for Bulimia Nervosa in both the KSADS-COMP and BN_SI_, over time by ***(A)*** caregiver-report and ***(B)*** youth-report.

#### Youth-report

Across all time points, 100 youth met the BN_SI_ criteria, while 90 youth met the KSADS-COMP criteria for BN. Jaccard indices ranged from 0.88 to 0.91, indicating high overlap between categorizations (see **Table 7**; **Figure 3B** displays frequency over time). Similar to the caregiver report, the BN_SI_ criteria identified additional participants who did not meet KSADS-COMP criteria.

### BED

#### Caregiver-report

Across all time points, 287 youth met the BED_SI_ criteria, while 276 youth met the KSADS-COMP criteria for BED. Jaccard indices ranged from 0.74 to 0.80, indicating that the KSADS-COMP and BED_SI_ classifications differed, although they yielded similar numbers of total classified youth (see **Table 8**; **Figure 4A** displays frequencies over time). This difference may be partially attributed to the methods, as youth who exhibited compensatory weight-control behaviors were excluded from classification as BED_SI_, consistent with *DSM-5* criteria. However, some of these youth were included in the KSADS-COMP definitions. Conversely, several youth who met BED_SI_ criteria were not identified by the KSADS-COMP algorithm, for unclear reasons.

**Table 8.**
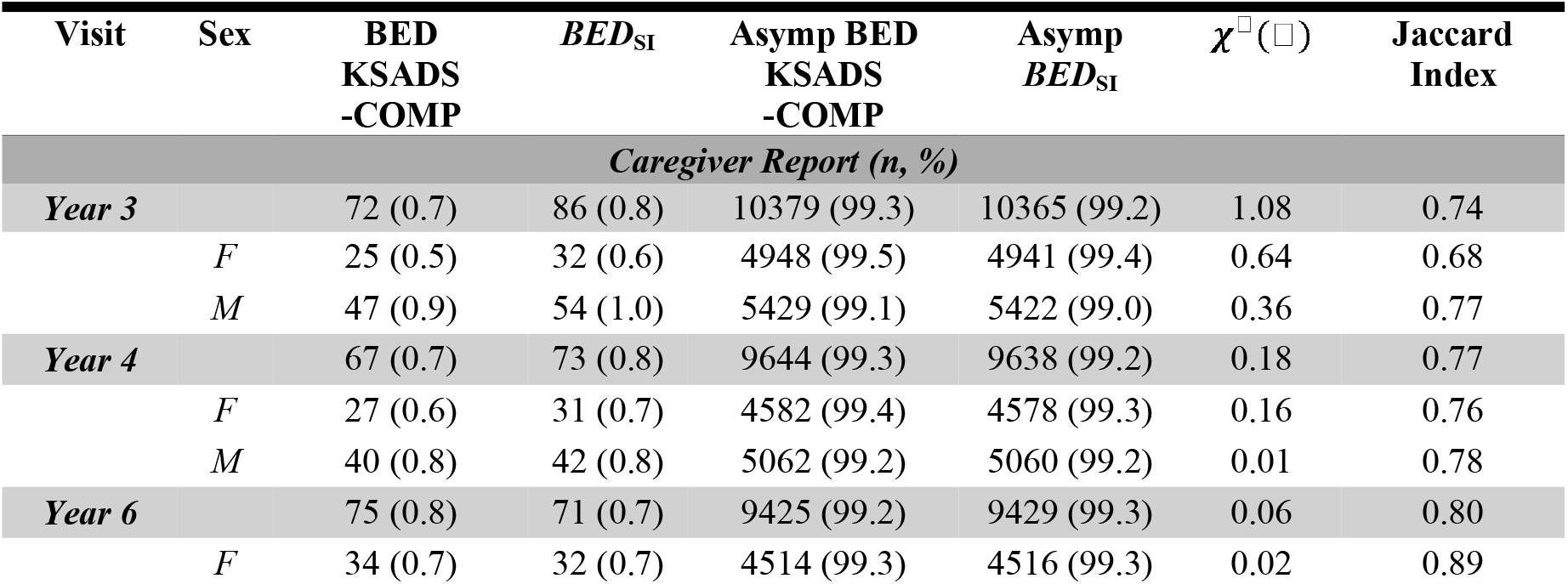
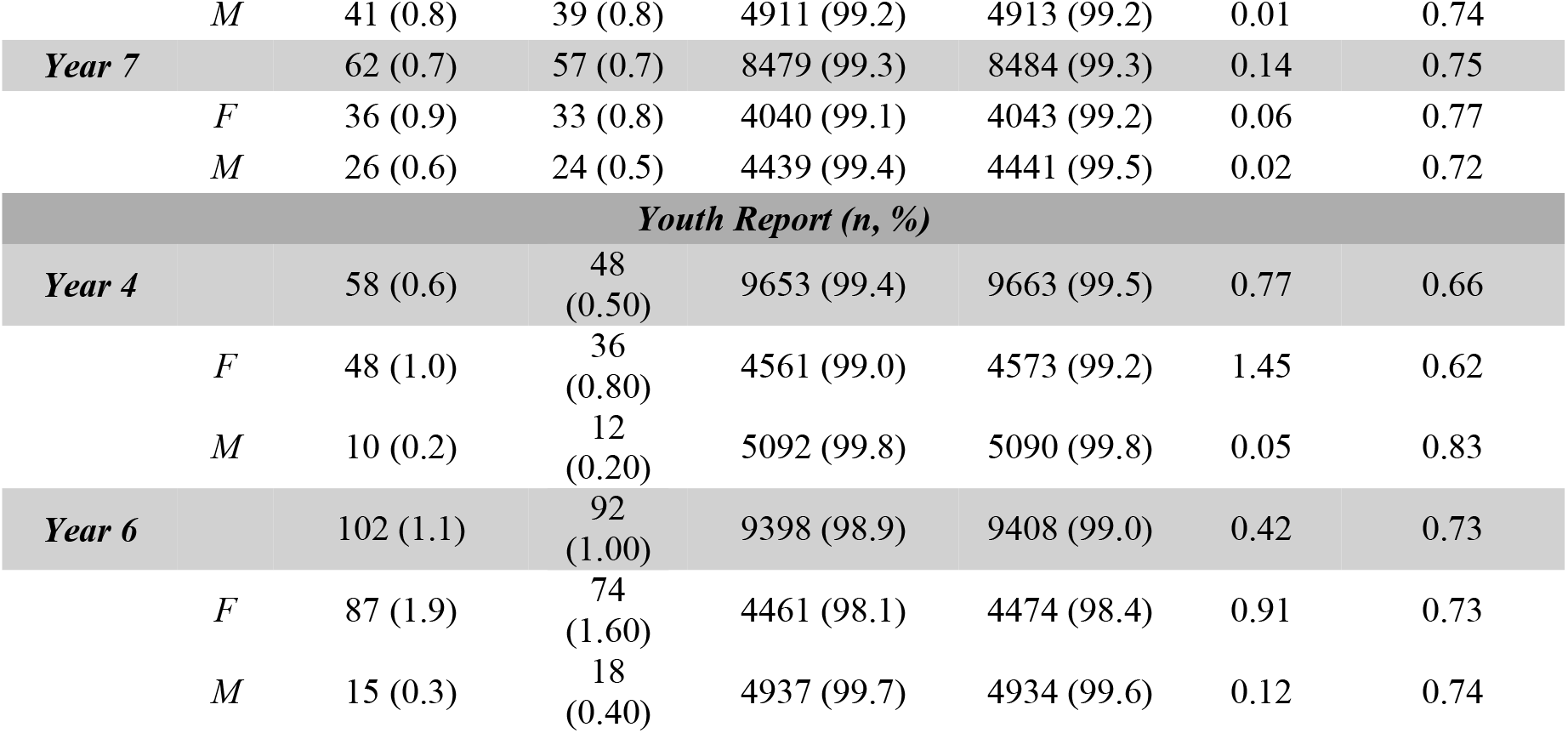
Number and proportion of youth who meet criteria for BED_SI_ compared to KSADS-COMP BED criteria. Data are presented as number of individuals meeting criteria and relative proportions in ABCD sample (%). BED_SI_ reflects youth who met the criteria for the Binge Eating Disorder Symptom Index. “Asymp” or “Asymptomatic” reflects youth who did not meet the criteria. Jaccard indices indicate the proportion of overlap in youth classified by both methods (KSADS-COMP vs. BED_SI_). Chi-squared statistics reflect tests of proportions for the KSADS-COMP vs. BED_SI_ at each time point, stratified by sex, with 1 degree of freedom and Yates continuity correction applied where possible. F = female; M = male; *** = p < 0.001; ** = p < 0.01; * = p < 0.05.

| **Visit** | **Sex** | **BED KSADS -COMP** | ***BED*<sub>SI</sub>** | **Asymp BED KSADS -COMP** | **Asymp *BED*<sub>SI</sub>** | ***χ*□(□)** | **Jaccard Index** |
|---|---|---|---|---|---|---|---|
| ***Caregiver Report (n, %)*** |  |  |  |  |  |  |  |
| ***Year 3*** |  | 72 (0.7) | 86 (0.8) | 10379 (99.3) | 10365 (99.2) | 1.08 | 0.74 |
|  | *F* | 25 (0.5) | 32 (0.6) | 4948 (99.5) | 4941 (99.4) | 0.64 | 0.68 |
|  | *M* | 47 (0.9) | 54 (1.0) | 5429 (99.1) | 5422 (99.0) | 0.36 | 0.77 |
| ***Year 4*** |  | 67 (0.7) | 73 (0.8) | 9644 (99.3) | 9638 (99.2) | 0.18 | 0.77 |
|  | *F* | 27 (0.6) | 31 (0.7) | 4582 (99.4) | 4578 (99.3) | 0.16 | 0.76 |
|  | *M* | 40 (0.8) | 42 (0.8) | 5062 (99.2) | 5060 (99.2) | 0.01 | 0.78 |
| ***Year 6*** |  | 75 (0.8) | 71 (0.7) | 9425 (99.2) | 9429 (99.3) | 0.06 | 0.80 |
|  | *F* | 34 (0.7) | 32 (0.7) | 4514 (99.3) | 4516 (99.3) | 0.02 | 0.89 |
|  | *M* | 41 (0.8) | 39 (0.8) | 4911 (99.2) | 4913 (99.2) | 0.01 | 0.74 |
| ***Year 7*** |  | 62 (0.7) | 57 (0.7) | 8479 (99.3) | 8484 (99.3) | 0.14 | 0.75 |
|  | *F* | 36 (0.9) | 33 (0.8) | 4040 (99.1) | 4043 (99.2) | 0.06 | 0.77 |
|  | *M* | 26 (0.6) | 24 (0.5) | 4439 (99.4) | 4441 (99.5) | 0.02 | 0.72 |
| ***Youth Report (n, %)*** |  |  |  |  |  |  |  |
| ***Year 4*** |  | 58 (0.6) | 48 (0.50) | 9653 (99.4) | 9663 (99.5) | 0.77 | 0.66 |
|  | *F* | 48 (1.0) | 36 (0.80) | 4561 (99.0) | 4573 (99.2) | 1.45 | 0.62 |
|  | *M* | 10 (0.2) | 12 (0.20) | 5092 (99.8) | 5090 (99.8) | 0.05 | 0.83 |
| ***Year 6*** |  | 102 (1.1) | 92 (1.00) | 9398 (98.9) | 9408 (99.0) | 0.42 | 0.73 |
|  | *F* | 87 (1.9) | 74 (1.60) | 4461 (98.1) | 4474 (98.4) | 0.91 | 0.73 |
|  | *M* | 15 (0.3) | 18 (0.40) | 4937 (99.7) | 4934 (99.6) | 0.12 | 0.74 |

**Figure 4.**
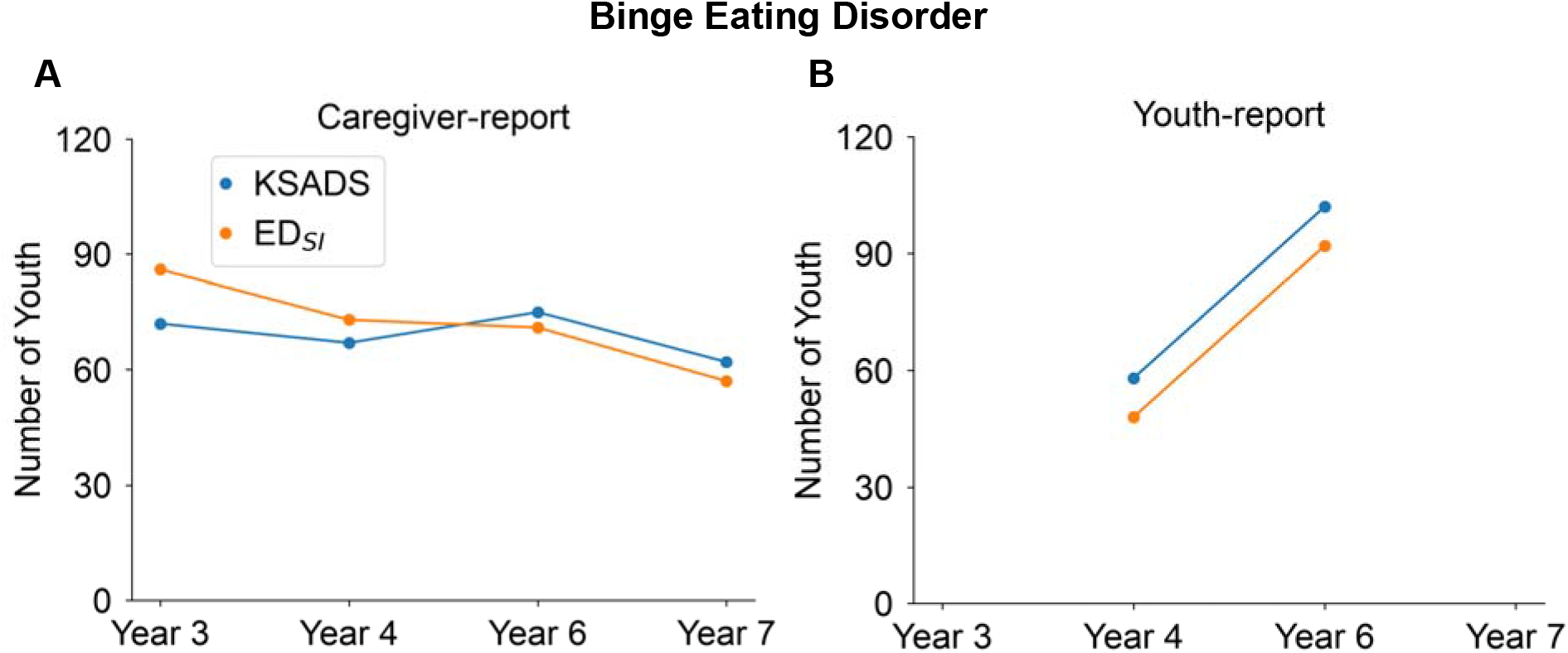
Number of youth who meet criteria for Binge Eating Disorder in both the KSADS-COMP and BED_SI_, over time by ***(A)*** caregiver-report and ***(B)*** youth-report.

#### Youth-report

Across all time points, 140 youth met BED_SI_ criteria, while 160 met KSADS-COMP criteria. Jaccard indices ranged from 0.66 to 0.73, indicating moderate to high overlap (see **Table 8**; **Figure 4B** displays frequency over time).

### OSFED

#### Caregiver-report

Across all time points, 3,219 youth met criteria for OSFED_SI_ (total across all sub-categories), while 100 youth met the KSADS-COMP criteria for OSFED (see **Table 9** for counts and frequencies separated by OSFED_SI_ sub-category; **Figure 5** displays frequency over time). Specifically, OSFED-AN-BP/BN_SI_ and OSFED-BED_SI_ captured significantly more youth than KSADS-COMP, accounting for heterogeneous cases of ED symptomatology. Low Jaccard indices (ranging from 0.02 to 0.19) indicated that ED_SI_ criteria generally do not capture the same youth as the KSADS-COMP.

**Table 9.**
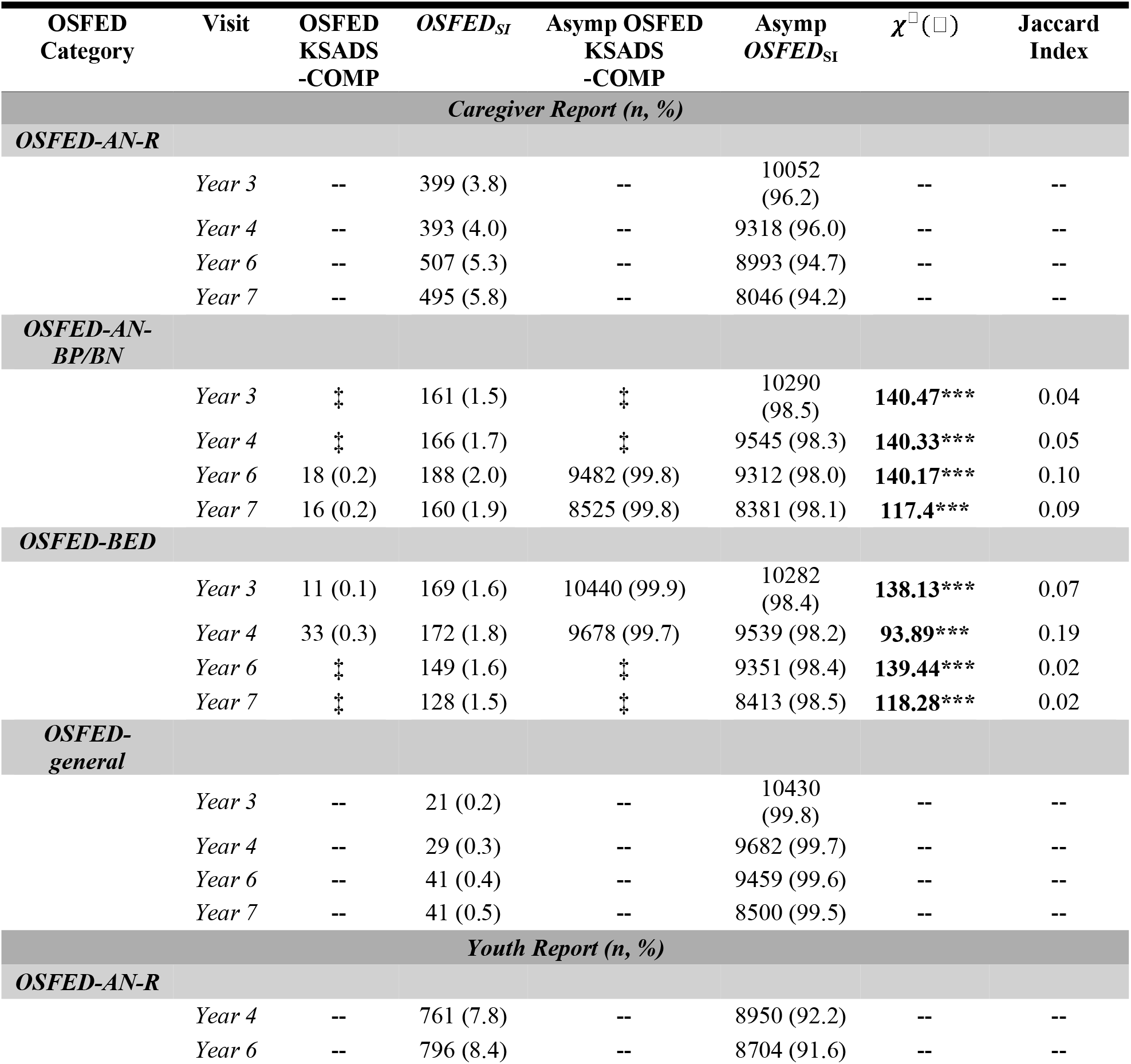
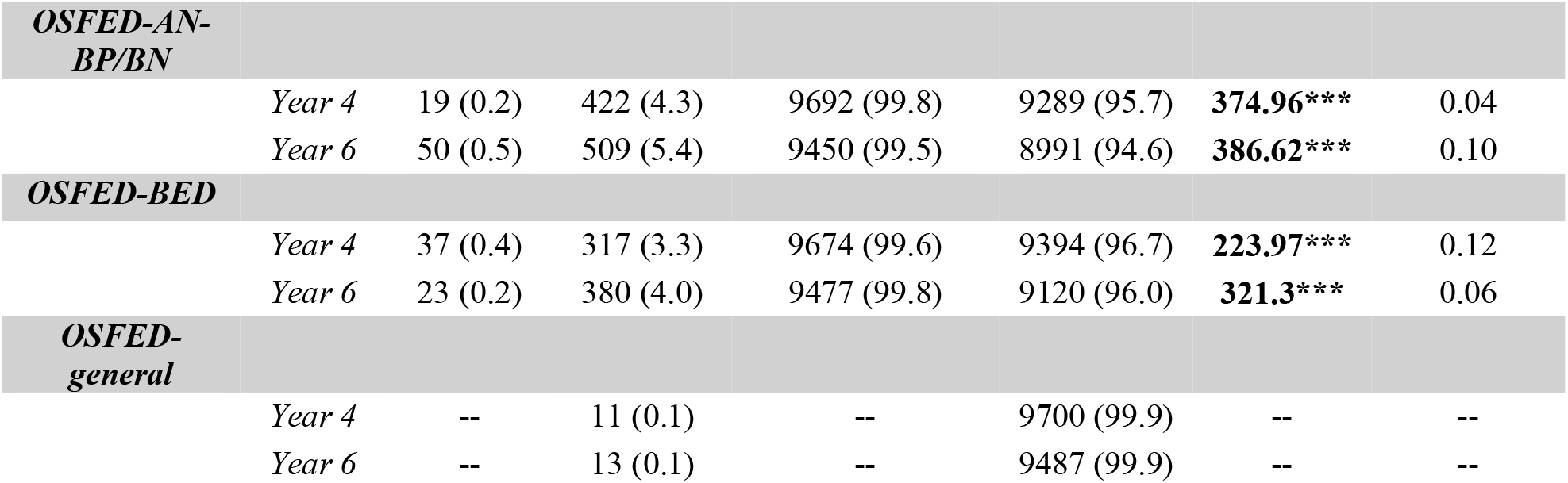
Number and proportion of youth who meet criteria for OSFED_SI_ compared to KSADS-COMP OSFED criteria. Data are presented as number of individuals meeting criteria and relative proportions in ABCD sample (%). OSFED_SI_ reflects youth who met the criteria for the Other Specified Feeding or Eating Disorder Symptom Index. “Asymp” or “Asymptomatic” reflects youth who did not meet the criteria. Chi-squared statistics reflect tests of proportions for the KSADS-COMP vs. OSFED_SI_ at each time point when comparisons were available, with 1 degree of freedom and Yates continuity correction applied where possible. Jaccard indices indicate the proportion of overlap in youth classified by both methods. *** = p < 0.001; ** = p < 0.01; * = p < 0. 05. Results stratified by sex can be found in Table S5. ‡ Reporting standards not met.

| OSFED Category | Visit | OSFED KSADS-COMP | *OSFED<sub>SI</sub>* | Asymp OSFED KSADS-COMP | Asymp *OSFED*<sub>SI</sub> | *χ*<sup>[illegible]</sup>([illegible]) | Jaccard Index |
|---|---|---|---|---|---|---|---|
| *Caregiver Report (n, %)* |  |  |  |  |  |  |  |
| ***OSFED-AN-R*** |  |  |  |  |  |  |  |
|  | *Year 3* | -- | 399 (3.8) | -- | 10052 (96.2) | -- | -- |
|  | *Year 4* | -- | 393 (4.0) | -- | 9318 (96.0) | -- | -- |
|  | *Year 6* | -- | 507 (5.3) | -- | 8993 (94.7) | -- | -- |
|  | *Year 7* | -- | 495 (5.8) | -- | 8046 (94.2) | -- | -- |
| ***OSFED-AN-BP/BN*** |  |  |  |  |  |  |  |
|  | *Year 3* | ‡ | 161 (1.5) | ‡ | 10290 (98.5) | **140.47\*\*\*** | 0.04 |
|  | *Year 4* | ‡ | 166 (1.7) | ‡ | 9545 (98.3) | **140.33\*\*\*** | 0.05 |
|  | *Year 6* | 18 (0.2) | 188 (2.0) | 9482 (99.8) | 9312 (98.0) | **140.17\*\*\*** | 0.10 |
|  | *Year 7* | 16 (0.2) | 160 (1.9) | 8525 (99.8) | 8381 (98.1) | **117.4\*\*\*** | 0.09 |
| ***OSFED-BED*** |  |  |  |  |  |  |  |
|  | *Year 3* | 11 (0.1) | 169 (1.6) | 10440 (99.9) | 10282 (98.4) | **138.13\*\*\*** | 0.07 |
|  | *Year 4* | 33 (0.3) | 172 (1.8) | 9678 (99.7) | 9539 (98.2) | **93.89\*\*\*** | 0.19 |
|  | *Year 6* | ‡ | 149 (1.6) | ‡ | 9351 (98.4) | **139.44\*\*\*** | 0.02 |
|  | *Year 7* | ‡ | 128 (1.5) | ‡ | 8413 (98.5) | **118.28\*\*\*** | 0.02 |
| ***OSFED-general*** |  |  |  |  |  |  |  |
|  | *Year 3* | -- | 21 (0.2) | -- | 10430 (99.8) | -- | -- |
|  | *Year 4* | -- | 29 (0.3) | -- | 9682 (99.7) | -- | -- |
|  | *Year 6* | -- | 41 (0.4) | -- | 9459 (99.6) | -- | -- |
|  | *Year 7* | -- | 41 (0.5) | -- | 8500 (99.5) | -- | -- |
| *Youth Report (n, %)* |  |  |  |  |  |  |  |
| ***OSFED-AN-R*** |  |  |  |  |  |  |  |
|  | *Year 4* | -- | 761 (7.8) | -- | 8950 (92.2) | -- | -- |
|  | *Year 6* | -- | 796 (8.4) | -- | 8704 (91.6) | -- | -- |
| ***OSFED-AN-BP/BN*** |  |  |  |  |  |  |  |
|  | *Year 4* | 19 (0.2) | 422 (4.3) | 9692 (99.8) | 9289 (95.7) | **374.96\*\*\*** | 0.04 |
|  | *Year 6* | 50 (0.5) | 509 (5.4) | 9450 (99.5) | 8991 (94.6) | **386.62\*\*\*** | 0.10 |
| ***OSFED-BED*** |  |  |  |  |  |  |  |
|  | *Year 4* | 37 (0.4) | 317 (3.3) | 9674 (99.6) | 9394 (96.7) | **223.97\*\*\*** | 0.12 |
|  | *Year 6* | 23 (0.2) | 380 (4.0) | 9477 (99.8) | 9120 (96.0) | **321.3\*\*\*** | 0.06 |
| ***OSFED-general*** |  |  |  |  |  |  |  |
|  | *Year 4* | -- | 11 (0.1) | -- | 9700 (99.9) | -- | -- |
|  | *Year 6* | -- | 13 (0.1) | -- | 9487 (99.9) | -- | -- |

**Figure 5.**
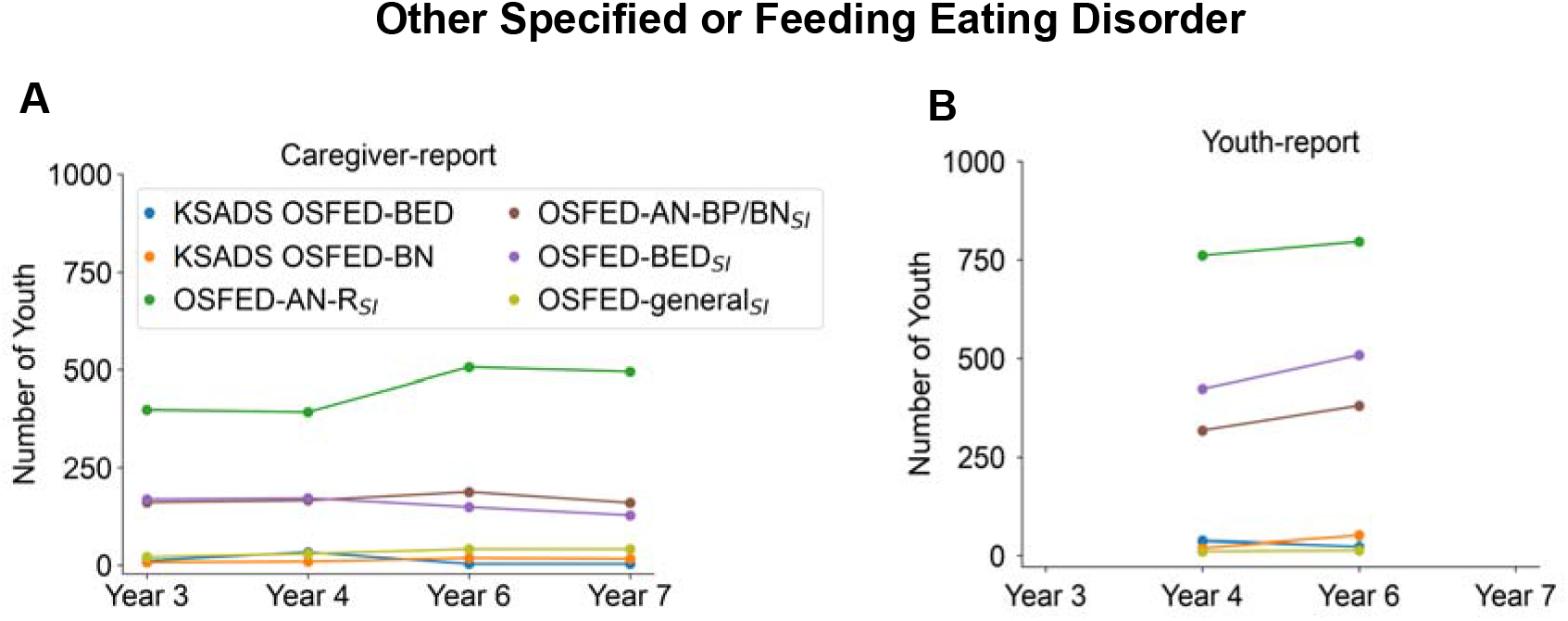
Number of youth who meet criteria for Other Specified Feeding or Eating Disorder (OSFED) in both the KSADS-COMP and OSFED_SI_, over time by ***(A)*** caregiver-report and ***(B)*** youth-report.

#### Youth-report

Across all time points, 3,209 youth met the OSFED_SI_ criteria, while 129 youth met the KSADS-COMP criteria for OSFED. Jaccard indices ranged from 0.06 to 0.12, indicating minimal overlap, similar to the caregiver-report (see **Table 9** for counts and frequencies by OSFED_SI_ subcategory; **Figure 5** shows frequency over time).

Counts and frequencies broken down by sex are included in Supplementary Materials (Table S5).

## DISCUSSION

The current manuscript presents a revised categorization strategy for utilizing KSADS-COMP ED item-level data to generate *DSM-5*-aligned ED_SI_ with the ABCD Study data.

Compared to KSADS-COMP-defined categories, Jaccard indices revealed noticeable differences in the number of youth captured across methods, particularly for AN_SI_ and OSFED_SI_; the differences for BN_SI_ and BED_SI_ were less divergent. Overlap among OSFED categories was more difficult to gauge, given that some of the distinct definitions provided by ED_SI_ are not captured by KSADS-COMP. Given the complexity and heterogeneity of ED presentations, transparent phenotyping is critical. Currently, it is unknown how KSADS-COMP categories are defined. Therefore, we provided transparent ED_SI_ definitions to capture youth who endorsed cognitive and behavioral signs and symptoms of AN, atypical AN, BN, and BED, as well as clinically relevant subtypes (e.g., AN-BP, AN-R) and subthreshold presentations (e.g., OSFED). Therefore, this new ED_SI_ categorization equips researchers with a structured approach to pursue ED-related scientific questions.

A primary aim of this manuscript was to compare the youth identified by ED_SI_ to those captured by the KSADS-COMP ED module scoring algorithm. At the aggregate level, differences were most pronounced for AN-spectrum presentations: the ED_SI_ identified virtually no cases of AN-BP_SI_ and substantially fewer cases of AN-R_SI_ and AAN_SI_ than the KSADS-COMP. Across all time points, relatively few youth met criteria for AN_SI_ regardless of reporter, reflecting lower than expected numbers based on broader epidemiological estimates (AN prevalence ∼1%) (Udo & Grilo, 2019). The requirement that youth be objectively underweight may be the most plausible explanation for these low counts. Indeed, substantially more assessment instances were classified as AAN-R_SI_, for which low weight was not required, including 222 caregiver-reported and 150 youth-reported instances across time points. It is possible that more youth would have met these criteria but lacked sufficiently reliable BMI data due to remote assessments or errors in height and weight measurements, resulting in biologically implausible values. Youth who met all other AN_SI_ criteria but lacked usable BMI data were therefore classified in the AN-wcu_SI_ category (n_caregiver_=109; n_youth_=40 across timepoints and subtypes). Given these data availability constraints and the small numbers within individual subtypes, combining AN_SI_, AAN_SI_, and AN-wcu_SI_ presentations may provide researchers with a more analytically useful measure for examining AN-spectrum phenotypes.

By contrast, the ED_SI_ and KSADS-COMP produced more similar aggregate counts for BN and BED. The prevalence of BN_SI_ and BED_SI_ was also more consistent with epidemiological estimates from pediatric and broader samples (Kjeldbjerg & Clausen, 2023). For example, at the 6-year follow-up, when participants were approximately 15–17 years old, 1.2% of females and 0.3% of males met criteria for BN_SI_. These estimates are comparable to previously reported point-prevalence estimates of 1.5% among females and 0.1% among males (Galmiche et al., 2019). Although BED typically has a later mean age of onset of approximately 23 years (Galmiche et al., 2019), youth-reported BED_SI_ was already identified in 1.6% of females and 0.4% of males at 6-year follow-up. These estimates align with previously reported point-prevalence estimates across age groups of 2.3% and 0.3% among females and males, respectively (Galmiche et al., 2019). Collectively, these findings suggest that the ED_SI_ definitions capture an overall burden of binge-eating-spectrum symptomatology broadly consistent with prior epidemiological findings.

The ED_SI_ also identified a substantial number of participants with OSFED_SI_ across four groups: youth with subthreshold AN-R, AN-BP or BN behaviors or BED; and youth with binge-eating or compensatory behaviors who did not meet criteria for another defined category. Direct comparison with KSADS-COMP was limited because several corresponding OSFED subtypes, namely OSFED-AN-R and OSFED-AN-BP/BN, were not defined in KSADS-COMP. Although these research-oriented OSFED_SI_ presentations do not correspond directly to the specific OSFED categories described in the *DSM-5*, they were intentionally designed to capture a broad range of transdiagnostic ED symptoms. Identifying these presentations may facilitate research on early or subthreshold symptoms that could precede the development of full-threshold EDs or be associated with clinically relevant comorbidities.

As the ABCD Study cohort enters the peak window for ED onset in mid-to-late adolescence (Hudson et al., 2007; Solmi et al., 2022), we anticipate that full-threshold ED_SI_ counts, particularly AN_SI_, will rise in future data releases. In the meantime, the comparatively higher yield of AAN_SI_, AN-wcu_SI_, and OSFED-AN-R_SI_ cases offers a valuable starting point for prospective designs aimed at identifying AN-like phenotypes and premorbid risk factors before youth cross the full diagnostic thresholds. We therefore encourage researchers to incorporate all four of these categories (i.e. AN_SI,_ AAN_SI_, AN-wcu_SI_, and OSFED-AN-R_SI_) when studying AN-related phenotypes, as this category may capture youth on similar risk trajectories showing cognitive/behavioral signs and symptoms consistent with those with AN_SI_ who have significantly low weight for their developmental stage. Subthreshold ED presentations also carry a degree of functional impairment comparable to full-threshold disorders (Fairweather-Schmidt & Wade, 2014; Stice et al., 2009) and most often precede them (Solmi et al., 2024). Thus, OSFED_SI_ categories may help identify youth who could be particularly informative for investigating premorbid risk factors within the ABCD Study, and may reflect earlier presentations of EDs, providing valuable insight for informing early intervention and prevention efforts.

Importantly, although categorization counts appeared similar across methods, Jaccard indices revealed substantial disagreement in which individual youth met criteria for specific categories (e.g., AN, BN, BED). Agreement was highest for BN_SI_ (Jaccard indices of 0.88-0.95 for caregiver-report and 0.88-0.91 for youth-report), moderate for BED_SI_ (0.66-0.80), and lowest for AN_SI_/AAN_SI_ (0.00-0.35) and OSFED_SI_ (0.02-0.19). Thus, similar aggregate prevalence estimates did not necessarily reflect agreement at the individual level. Concordance was greatest for BN_SI_ and BED_SI,_ which may reflect greater reliance on visible, outward behaviors and relatively frequent symptoms.

Regarding BED, disagreement may stem in part from differences in how the *DSM-5* criteria were operationalized. Our empirically supported operationalization explicitly excluded youth who engaged in compensatory behaviors from meeting the criteria for BED (Abber et al., 2026). Individual-level examination of youth classified as having BED by KSADS-COMP suggested that its algorithm may not have consistently applied this exclusion, potentially accounting for some of the observed discordance. For AN_SI_ and AAN_SI_, lower concordance may instead reflect differences in how weight and symptom criteria are operationalized. When available, our AN_SI_/AAN_SI_ classifications incorporated objectively measured height and weight, whereas the KSADS-COMP ED module relied on caregiver-or youth-reported anthropometric estimates. The approaches also differed in how they operationalized the “fear of weight gain” criterion. Our AN_SI_/AAN_SI_/AN-wcu_SI_ criteria used a more liberal threshold (e.g., ≥ 1 symptom on a 0–4 scale), whereas the KSADS-COMP ED algorithm imposed a more stringent cutoff (e.g., >3). However, sensitivity analyses indicated that the more liberal threshold for “fear of weight gain” did not account for the observed differences between methods. Together, these findings suggest that discordance in AN-related classification may be driven more by differences in the measurement and operationalization of each criteria than by differences in overall case frequency.

Even more pronounced discrepancies emerged for OSFED_SI_ and KSADS-COMP OSFED groups, showing consistently low Jaccard indices, suggesting that the two methods largely identified different youth with ED symptoms. This was expected to some degree, given that our OSFED_SI_ definitions were intentionally constructed to be inclusive of any youth who endorsed some level of ED symptoms, including OSFED-AN-R_SI_ and OSFED-AN-BP/BN_SI_ categories that had no KSADS-COMP analogs. The larger pool of youth with OSFED_SI_ may also help researchers more comprehensively identify youth with subthreshold ED symptomatology in the ABCD Study dataset. In turn, this could help benchmark proxy prevalence estimates of OSFED in non-treatment-seeking samples, given that existing OSFED prevalence estimates have largely been derived from clinical samples (Solmi et al., 2024).

Moreover, regardless of classification method, groupings from youth-vs. caregiver-reports differed substantially at the 4-and 6-year follow-ups (∼ages 13–17 years). This is consistent with previous findings of informant differences in the ABCD study’s mental health measures (Baskin-Sommers et al., 2026; Costello et al., 2011; Karcher & Barch, 2021). Youth-reported cases for AAN, BN, BED, and OSFED consistently exceeded caregiver-reports across both ED_SI_ and KSADS-COMP. For example, at 6-year follow-up, youth-reported AAN and BN instances were approximately 1.6-fold and 2-fold higher than caregiver reports, respectively.

While a full exploration of these informant discrepancies is beyond the scope of this manuscript, these patterns underscore that ED_SI_ surveillance in the ABCD Study should carefully consider data from both reporters depending on the research question, where inclusion of both reporters could provide complementary and/or additive information.

### Toward more reproducible ED research in ABCD

It is important to note that a precise item-level comparison between ED_SI_ and KSADS-COMP across all included ED categories is not possible, as the KSADS-COMP scoring algorithm is not publicly available. Ultimately, the misalignments between these two methods underscores the critical need for reproducible, standardized, and explicitly operationalized criteria, which the ED_SI_ directly addresses. This is particularly relevant given recently documented inconsistencies among ABCD data including neuroimaging, much of which has been attributed to heterogeneous, non-transparent case definitions across studies (Martin et al., 2026). By providing fully transparent, item-level criteria and openly available code, ED_SI_ offers a common methodological foundation from which ABCD study researchers can consistently derive and report on ED symptoms. More broadly, the longitudinal design and multimodal depth of the ABCD Study make it uniquely positioned to advance understanding of the developmental origins of EDs. The comparatively higher numbers of youth with AAN_SI_ and OSFED-AN-R_SI_ relative to full-threshold AN_SI_ is particularly valuable in this context, as ABCD’s temporal design will allow researchers to pinpoint when and for whom subsequent ED symptomatology evolves (Solmi et al., 2024).

This work needs to be interpreted considering certain limitations. The KSADS-COMP does not assess all *DSM-5* ED categories, including avoidant restrictive food intake disorder (ARFID), pica, and rumination disorder. Thus, ED_SI_ cannot capture the full spectrum of adolescent eating pathology. ED_SI_ also approximates rather than fully replicates *DSM-5* criteria, given constraints to specific items available in the KSADS-COMP, including the collection of objective BMI data to distinguish between AN subtypes and other ED categories, as described above. The KSADS-COMP scoring algorithm is not publicly documented; thus, our explanations for discrepancies between ED_SI_ and KSADS-COMP classifications are inferential. Analyses presented here were restricted to KSADS-COMP Version 2 (corresponding to 3-year follow-up onward), given challenges in curating data across KSADS-COMP versions in current releases; however, corrections are expected in the 7.1 patch release. Finally, ED_SI_ reflects youth-or caregiver-reported rather than clinician-assessed ED symptomatology and thus should not be interpreted as formal diagnoses.

## CONCLUSION

ED_SI_ provides the research community with a transparent, reproducible, and developmentally sensitive tool for characterizing ED symptomatology in one of the largest, longitudinal pediatric cohorts available. As the ABCD Study cohort ages into the peak window for ED onset, we anticipate that ED_SI_ will support studies capable of identifying key pathways across neurobiological, metabolic, genetic, and environmental domains that give rise to and maintain disordered eating in adolescence.

## Data Availability

The ABCD data used in this report came from ABCD Study data release 7.0, which can be accessed through the NIH Brain Development Cohorts (NBDC) platform: https://www.nbdc-datahub.org/.

## ACKNOWLEDGMENTS/FUNDING

The Adolescent Brain Cognitive Development Study (ABCD Study^®^) is supported by the National Institutes of Health and National Institute on Drug Abuse and additional federal partners under award numbers U01DA041048, U01DA050989, U01DA051016, U01DA041022, U01DA051018, U01DA051037, U01DA050987, U01DA041174, U01DA041106, U01DA041117, U01DA041028, U01DA041134, U01DA050988, U01DA051039, U01DA041156, U01DA041025, U01DA041120, U01DA051038, U01DA041148, U01DA041093, U01DA041089, U24DA041123, U24DA041147. A full list of supporters is available at https://abcdstudy.org/federal-partners/. A listing of participating sites and a complete listing of the study investigators can be found at https://abcdstudy.org/principal-investigators/. The ABCD Study^®^ consortium investigators designed and implemented the study and/or provided data but did not necessarily participate in analysis or writing of this report. This manuscript reflects the views of the authors and may not reflect the opinions or views of the NIH or ABCD Consortium investigators. The ABCD data repository grows and changes over time. The ABCD data used in this report came from ABCD Study data release 7.0, which can be accessed through the NIH Brain Development Cohorts (NBDC) platform: https://www.nbdc-datahub.org/.

Additional support for this work was made possible by NIDDK (K01 DK135847; awarded to S.A.) and NIMH (R00MH132866 awarded to C.M., K23MH137567 awarded to K.H.).

## Data and code availability

The data used in this manuscript are from the ABCD Study data release 7.0 (DOI: https://doi.org/10.82525/8f3w-5260). Importantly, the KSADS-COMP underwent an update after the two-year follow-up of the ABCD Study, including changes to the questions, branching logic, and scoring algorithms. Therefore, for the purpose of this manuscript and to bypass challenges in ongoing harmonization efforts across versions for future releases, we report only on KSADS-COMP Version 2 (e.g., the three-year follow-up onwards). Accordingly, estimates presented in this manuscript should not be interpreted as prevalence rates, as these numbers may change once data harmonization is complete. Code to generate the ED_SI_ groups will be available upon publication of the manuscript.

## Author contributions

SA, CM, DB, BM, KR, KH, and KG wrote the manuscript. KR, CM, SA conceptualized the research idea. CM and SA interpreted the results. All authors contributed to the manuscript review and editing.

