## Supplementary Material for "Mapping disordered eating symptoms in the ABCD Study through the Eating Disorder Symptom Index"

**Table S1. OSFED Anorexia Nervosa Restricting Symptom Index Criteria (OSFED-AN-R_SI_)**

*Case 1*

A. Fear of weight gain

B. Self-worth tied to weight

~~C. Weight status~~

D. Sufficient frequency

~~E. Sufficient duration~~

F. Restriction (a), AND No endorsement of binge/purge symptoms (b)

*Case 2*

A. Fear of weight gain

~~B. Self-worth tied to weight~~

~~C. Weight status~~

D. Sufficient frequency

E. Sufficient duration

F. Restriction (a), AND No endorsement of binge/purge symptoms (b)

*Case 3*

A. Fear of weight gain

~~B. Self-worth tied to weight~~

~~C. Weight status~~

D. Sufficient frequency

~~E. Sufficient duration~~

F. Restriction (a), AND No endorsement of binge/purge symptoms (b)

*Case 4*

A. Fear of weight gain

~~B. Self-worth tied to weight~~

~~C. Weight status~~

~~D. Sufficient frequency~~

~~E. Sufficient duration~~

F. Restriction (a), AND No endorsement of binge/purge symptoms (b)

*Case 5*

~~A. Fear of weight gain~~

~~B. Self-worth tied to weight~~

~~C. Weight status~~

~~D. Sufficient frequency~~

~~E. Sufficient duration~~

F. Restriction (a), AND No endorsement of binge/purge symptoms (b)

*Note. Cases list full index criteria for AN-R-_SI_, with specific criteria crossed out that participants may not have met. Participants who met the remaining criteria for any listed case met criteria for AN-R-OSFED-_SI_. Questions asked in the KSADS-COMP participant ED module corresponding to each criterion can be found in Table 1 in the Methods section.*

**Table S2. OSFED Anorexia-Binge/Purge / Bulimia Nervosa Symptom Index (OSFED-AN-BP/BN_SI_)**

*BN-related Case 1*

A. Binge eating

B. Compensatory weight control

C. Self-worth tied to weight

D. Sufficient frequency of binge eating

~~E. Sufficient duration of binge eating~~

F. Sufficient frequency of compensatory weight control

G. Sufficient duration of compensatory weight control

*BN-related Case 2*

A. Binge eating

B. Compensatory weight control

C. Self-worth tied to weight

D. Sufficient frequency of binge eating

E. Sufficient duration of binge eating

F. Sufficient frequency of compensatory weight control

~~G. Sufficient duration of compensatory weight control~~

*BN-related Case 3*

A. Binge eating

B. Compensatory weight control

C. Self-worth tied to weight

D. Sufficient frequency of binge eating

~~E. Sufficient duration of binge eating~~

F. Sufficient frequency of compensatory weight control

~~G. Sufficient duration of compensatory weight control~~

*BN-related Case 4*

A. Binge eating

B. Compensatory weight control

~~C. Self-worth tied to weight~~

D. Sufficient frequency of binge eating

E. Sufficient duration of binge eating

F. Sufficient frequency of compensatory weight control

G. Sufficient duration of compensatory weight control

*BN-related Case 5*

A. Binge eating

B. Compensatory weight control

~~C. Self-worth tied to weight~~

D. Sufficient frequency of binge eating

~~E. Sufficient duration of binge eating~~

F. Sufficient frequency of compensatory weight control

~~G. Sufficient duration of compensatory weight control~~

*BN-related Case 6*

A. Binge eating

B. Compensatory weight control

~~C. Self-worth tied to weight~~

~~D. Sufficient frequency of binge eating~~

~~E. Sufficient duration of binge eating~~

~~F. Sufficient frequency of compensatory weight control~~

~~G. Sufficient duration of compensatory weight control~~

*AN-BP-related Case 1*

A. Fear of weight gain

~~B. Self-worth tied to weight~~

~~C. Weight status~~

D. Sufficient frequency

E. Sufficient duration

G. Endorsement of binge/purge symptoms

*AN-BP-related Case 2*

A. Fear of weight gain

B. Self-worth tied to weight

~~C. Weight status~~

D. Sufficient frequency

~~E. Sufficient duration~~

G. Endorsement of binge/purge symptoms

*AN-BP-related Case 3*

~~A. Fear of weight gain~~

B. Self-worth tied to weight

~~C. Weight status~~

D. Sufficient frequency

E. Sufficient duration

G. Endorsement of binge/purge symptoms

*AN-BP-related Case 4*

~~A. Fear of weight gain~~

~~B. Self-worth tied to weight~~

~~C. Weight status~~

D. Sufficient frequency

E. Sufficient duration

G. Endorsement of binge/purge symptoms

*AN-BP-related Case 5*

~~A. Fear of weight gain~~

~~B. Self-worth tied to weight~~

~~C. Weight status~~

D. Sufficient frequency

~~E. Sufficient duration~~

G. Endorsement of binge/purge symptoms

*AN-BP-related Case 6*

~~A. Fear of weight gain~~

~~B. Self-worth tied to weight~~

~~C. Weight status~~

~~D. Sufficient frequency~~

~~E. Sufficient duration~~

G. Endorsement of binge/purge symptoms

*Note. AN-BP-related cases list full index criteria for AN-BP_SI_, with specific criteria crossed out that participants may not have met. Likewise, BN-related cases list full index criteria for BN_SI_, with specific criteria crossed out that participants may not have met. Participants who met remaining criteria for any listed case met criteria for OSFED-AN-BP/BN_SI_. Questions asked in the KSADS-COMP participant ED module corresponding to each criterion can be found in Tables 1 and 2 in the Methods section.*

**Table S3. OSFED Binge Eating Disorder Symptom Index (OSFED-BED_SI_)**

*Case 1*

A. Binge eating

~~B. Symptoms of binge eating (Must endorse at least 3 symptoms of options 1-7)~~

C. Distress while binge eating

D. Sufficient frequency of binge eating

E. Sufficient duration of binge eating

F. No compensatory weight-control

*Case 2*

A. Binge eating

B. Symptoms of binge eating (Must endorse at least 3 symptoms of options 1-7)

C. Distress while binge eating

D. Sufficient frequency of binge eating

~~E. Sufficient duration of binge eating~~

F. No compensatory weight-control

*Case 3*

A. Binge eating

B. Symptoms of binge eating (Must endorse at least 3 symptoms of options 1-7)

~~C. Distress while binge eating~~

D. Sufficient frequency of binge eating

E. Sufficient duration of binge eating

F. No compensatory weight-control

*Case 4*

A. Binge eating

B. Symptoms of binge eating (Must endorse at least *1 symptom* of options 1-7)

C. Distress while binge eating

D. Sufficient frequency of binge eating

E. Sufficient duration of binge eating

F. No compensatory weight-control

*Case 5*

A. Binge eating

B. Symptoms of binge eating (Must endorse at least *1 symptom* of options 1-7)

C. Distress while binge eating

D. Sufficient frequency of binge eating

~~E. Sufficient duration of binge eating~~

F. No compensatory weight-control

*Case 6*

A. Binge eating

B. Symptoms of binge eating (Must endorse at least 3 symptoms of options 1-7)

C. Distress while binge eating

~~D. Sufficient frequency of binge eating~~

E. Sufficient duration of binge eating

F. No compensatory weight-control

*Case 7*

A. Binge eating

B. Symptoms of binge eating (Must endorse at least *1 symptom* of options 1-7)

~~C. Distress while binge eating~~

D. Sufficient frequency of binge eating

~~E. Sufficient duration of binge eating~~

F. No compensatory weight-control

*Case 8*

A. Binge eating

B. Symptoms of binge eating (Must endorse at least *1 symptom* of options 1-7)

~~C. Distress while binge eating~~

~~D. Sufficient frequency of binge eating~~

~~E. Sufficient duration of binge eating~~

F. No compensatory weight-control

*Case 9*

A. Binge eating

~~B. Symptoms of binge eating (Must endorse at least 3 symptoms of options 1-7)~~

~~C. Distress while binge eating~~

~~D. Sufficient frequency of binge eating~~

~~E. Sufficient duration of binge eating~~

F. No compensatory weight-control

*Case 10*

~~A. Binge eating~~

~~B. Symptoms of binge eating (Must endorse at least 3 symptoms of options 1-7)~~

~~C. Distress while binge eating~~

D. Sufficient frequency of binge eating

~~E. Sufficient duration of binge eating~~

F. No compensatory weight-control

*Note. Cases list full index criteria for BED_SI_, with specific criteria crossed out that participants may not have met. Participants who met remaining criteria for any listed case met criteria for OSFED-BED_SI_. Questions asked in the KSADS-COMP participant ED module corresponding to each criterion can be found in Table 3 in the Methods section.*

**Table S4. OSFED General Symptom Index (OSFED-general_SI_)**

*General Case 1*

A. Sufficient frequency of compensatory weight control

*General Case 2*

A. Sufficient duration of compensatory weight control

*Note. These two “general” cases were included to capture any additional ED symptomology presentations not already included in the previous OSFED_SI_ categories. Questions asked in the KSADS-COMP participant ED module corresponding to each criterion can be found in Table 1 in the Methods section.*

| Table S5. Number and proportion of youth who meet criteria for OSFED_SI_ compared to KSADS-COMP OSFED criteria, stratified by sex. | | | | | | | | |
| --- | --- | --- | --- | --- | --- | --- | --- | --- |
| OSFED Category | Visit | Sex | OSFED  KSADS  -COMP | *OSFED_SI_* | Asymptomatic OSFED  KSADS  -COMP | Asymptomatic  *OSFED*_SI_ | $\boldsymbol{\chi}^{\boldsymbol{2}}\boldsymbol{(1)}$ | Jaccard Index |
| *Caregiver Report (n, %)* | | | | | | | | |
| *OSFED-AN-R* |  |  |  |  |  |  |  |  |
|  | *Year 3* |  | -- | 399 (3.8) | -- | 10052 (96.2) | -- | -- |
|  |  | *F* | -- | 209 (4.2) | -- | 4764 (95.8) | -- | -- |
|  |  | *M* | -- | 190 (3.5) | -- | 5286 (96.5) | -- | -- |
|  | *Year 4* |  | -- | 393 (4.0) | -- | 9318 (96.0) | -- | -- |
|  |  | *F* | -- | 212 (4.6) | -- | 4397 (95.4) | -- | -- |
|  |  | *M* | -- | 181 (3.5) | -- | 4921 (96.5) | -- | -- |
|  | *Year 6* |  | -- | 507 (5.3) | -- | 8993 (94.7) | -- | -- |
|  |  | *F* | -- | 264 (5.8) | -- | 4284 (94.2) | -- | -- |
|  |  | *M* | -- | 243 (4.9) | -- | 4709 (95.1) | -- | -- |
|  | *Year 7* |  | -- | 495 (5.8) | -- | 8046 (94.2) | -- | -- |
|  |  | *F* | -- | 249 (6.1) | -- | 3827 (93.9) | -- | -- |
|  |  | *M* | -- | 246 (5.5) | -- | 4219 (94.5) | -- | -- |
| *OSFED-AN-BP/BN* |  |  |  |  |  |  |  |  |
|  | *Year 3* |  | ‡ | 161 (1.5) | ‡ | 10290 (98.5) | 140.47*** | 0.04 |
|  |  | *F* | ‡ | 77 (1.5) | ‡ | 4896 (98.5) | 67.15*** | 0.04 |
|  |  | *M* | ‡ | 84 (1.5) | ‡ | 5392 (98.5) | 71.49*** | 0.04 |
|  | *Year 4* |  | ‡ | 166 (1.7) | ‡ | 9545 (98.3) | 140.33*** | 0.05 |
|  |  | *F* | ‡ | 91 (2.0) | ‡ | 4518 (98.0) | 84.12*** | 0.02 |
|  |  | *M* | ‡ | 75 (1.5) | ‡ | 5027 (98.5) | 55.19*** | 0.09 |
|  | *Year 6* |  | 18 (0.2) | 188 (2.0) | 9482 (99.8) | 9312 (98.0) | 140.17*** | 0.10 |
|  |  | *F* | ‡ | 116 (2.6) | ‡ | 4432 (97.4) | 77.45*** | 0.13 |
|  |  | *M* | ‡ | 72 (1.5) | ‡ | 4880 (98.5) | 62.12*** | 0.04 |
|  | *Year 7* |  | 16 (0.2) | 160 (1.9) | 8525 (99.8) | 8381 (98.1) | 117.4*** | 0.09 |
|  |  | *F* | ‡ | 86 (2.1) | ‡ | 3990 (97.9) | 59.29*** | 0.10 |
|  |  | *M* | ‡ | 74 (1.7) | ‡ | 4391 (98.3) | 56.62*** | 0.07 |
| *OSFED-BED* |  |  |  |  |  |  |  |  |
|  | *Year 3* |  | 11 (0.1) | 169 (1.6) | 10440 (99.9) | 10282 (98.4) | 138.13*** | 0.07 |
|  |  | *F* | ‡ | 74 (1.5) | ‡ | 4899 (98.5) | 59*** | 0.07 |
|  |  | *M* | ‡ | 95 (1.7) | ‡ | 5381 (98.3) | 77.39*** | 0.06 |
|  | *Year 4* |  | 33 (0.3) | 172 (1.8) | 9678 (99.7) | 9539 (98.2) | 93.89*** | 0.19 |
|  |  | *F* | 17 (0.4) | 78 (1.7) | 4592 (99.6) | 4531 (98.3) | 38.29*** | 0.22 |
|  |  | *M* | 16 (0.3) | 94 (1.8) | 5086 (99.7) | 5008 (98.2) | 54.49*** | 0.17 |
|  | *Year 6* |  | ‡ | 149 (1.6) | ‡ | 9351 (98.4) | 139.44*** | 0.02 |
|  |  | *F* | ‡ | 81 (1.8) | ‡ | 4467 (98.2) | 73.98*** | 0.02 |
|  |  | *M* | ‡ | 68 (1.4) | ‡ | 4884 (98.6) | 63.57*** | 0.01 |
|  | *Year 7* |  | ‡ | 128 (1.5) | ‡ | 8413 (98.5) | 118.28*** | 0.02 |
|  |  | *F* | ‡ | 63 (1.5) | ‡ | 4013 (98.5) | 53.17*** | 0.05 |
|  |  | *M* | ‡ | 65 (1.5) | ‡ | 4400 (98.5) | 63.48** | 0.00 |
| *OSFED-*  *general* |  |  |  |  |  |  |  |  |
|  | *Year 3* |  | -- | 21 (0.2) | -- | 10430 (99.8) | -- | -- |
|  |  | *F* | -- | ‡ | -- | ‡ | -- | -- |
|  |  | *M* | -- | ‡ | -- | ‡ | -- | -- |
|  | *Year 4* |  | -- | 29 (0.3) | -- | 9682 (99.7) | -- | -- |
|  |  | *F* | -- | 11 (0.2) | -- | 4598 (99.8) | -- | -- |
|  |  | *M* | -- | 18 (0.4) | -- | 5084 (99.6) | -- | -- |
|  | *Year 6* |  | -- | 41 (0.4) | -- | 9459 (99.6) | -- | -- |
|  |  | *F* | -- | 19 (0.4) | -- | 4529 (99.6) | -- | -- |
|  |  | *M* | -- | 22 (0.4) | -- | 4930 (99.6) | -- | -- |
|  | *Year 7* |  | -- | 41 (0.5) | -- | 8500 (99.5) | -- | -- |
|  |  | *F* | -- | 18 (0.4) | -- | 4058 (99.6) | -- | -- |
|  |  | *M* | -- | 23 (0.5) | -- | 4442 (99.5) | -- | -- |
| *Youth Report (n, %)* | | | | | | | | |
| *OSFED-AN-R* |  |  |  |  |  |  |  |  |
|  | *Year 4* |  | -- | 761 (7.8) | -- | 8950 (92.2) | -- | -- |
|  |  | *F* | -- | 358 (7.8) | -- | 4251 (92.2) | -- | -- |
|  |  | *M* | -- | 403 (7.9) | -- | 4699 (92.1) | -- | -- |
|  | *Year 6* |  | -- | 796 (8.4) | -- | 8704 (91.6) | -- | -- |
|  |  | *F* | -- | 373 (8.2) | -- | 4175 (91.8) | -- | -- |
|  |  | *M* | -- | 423 (8.5) | -- | 4529 (91.5) | -- | -- |
| *OSFED-AN-BP/BN* |  |  |  |  |  |  |  |  |
|  | *Year 4* |  | 19 (0.2) | 422 (4.3) | 9692 (99.8) | 9289 (95.7) | **374.96***** | 0.04 |
|  |  | *F* | ‡ | 266 (5.8) | ‡ | 4343 (94.2) | **229.41***** | 0.06 |
|  |  | *M* | ‡ | 156 (3.1) | ‡ | 4946 (96.9) | **144.78***** | 0.02 |
|  | *Year 6* |  | 50 (0.5) | 509 (5.4) | 9450 (99.5) | 8991 (94.6) | **386.62***** | 0.10 |
|  |  | *F* | 37 (0.8) | 302 (6.6) | 4511 (99.2) | 4246 (93.4) | **213.55***** | 0.12 |
|  |  | *M* | 13 (0.3) | 207 (4.2) | 4939 (99.7) | 4745 (95.8) | **173.16***** | 0.06 |
| *OSFED-BED* |  |  |  |  |  |  |  |  |
|  | *Year 4* |  | 37 (0.4) | 317 (3.3) | 9674 (99.6) | 9394 (96.7) | **223.97***** | 0.12 |
|  |  | *F* | 27 (0.6) | 174 (3.8) | 4582 (99.4) | 4435 (96.2) | **108.41***** | 0.16 |
|  |  | *M* | 10 (0.2) | 143 (2.8) | 5092 (99.8) | 4959 (97.2) | **115.62***** | 0.07 |
|  | *Year 6* |  | 23 (0.2) | 380 (4.0) | 9477 (99.8) | 9120 (96.0) | **321.3***** | 0.06 |
|  |  | *F* | ‡ | 211 (4.6) | ‡ | 4337 (95.4) | **165.14***** | 0.09 |
|  |  | *M* | ‡ | 169 (3.4) | ‡ | 4783 (96.6) | **155.43***** | 0.03 |
| *OSFED-*  *general* |  |  |  |  |  |  |  |  |
|  | *Year 4* |  | -- | 11 (0.1) | -- | 9700 (99.9) | -- | -- |
|  |  | *F* | -- | ‡ | -- | ‡ | -- | -- |
|  |  | *M* | -- | ‡ | -- | ‡ | -- | -- |
|  | *Year 6* |  | -- | 13 (0.1) | -- | 9487 (99.9) | -- | -- |
|  |  | *F* | -- | ‡ | -- | ‡ | -- | -- |
|  |  | *M* | -- | ‡ | -- | ‡ | -- | -- |

*Note. Data are numbers and percentages (%). OSFED*_SI_ *reflects youth who met the criteria for the Other Specified Feeding or Eating Disorder Symptom Index. “Asymptomatic” reflects youth who did not meet the criteria. Chi-squared statistics reflect tests of proportions for the KSADS-COMP vs OSFED*_SI_ *at each time point when comparisons were available, stratified by sex, with 1 degree of freedom and Yates continuity correction applied where possible. Jaccard indices indicate the proportion of overlap in youth classified by both methods. F = female; M = male; *** = p < 0.001; ** = p < 0.01; * = p < 0.05. Results stratified by sex can be found in Table S5.*

*‡ Reporting standards not met.*
